# Decoding Clinical Biomarker Space of COVID-19: Exploring Matrix Factorization-based Feature Selection Methods

**DOI:** 10.1101/2021.07.07.21259699

**Authors:** Farshad Saberi-Movahed, Mahyar Mohammadifard, Adel Mehrpooya, Mohammad Rezaei-Ravari, Kamal Berahmand, Mehrdad Rostami, Saeed Karami, Mohammad Najafzadeh, Davood Hajinezhad, Mina Jamshidi, Farshid Abedi, Mahtab Mohammadifard, Elnaz Farbod, Farinaz Safavi, Mohammadreza Dorvash, Shahrzad Vahedi, Mahdi Eftekhari, Farid Saberi-Movahed, Iman Tavassoly

## Abstract

One of the most critical challenges in managing complex diseases like COVID-19 is to establish an intelligent triage system that can optimize the clinical decision-making at the time of a global pandemic. The clinical presentation and patients’ characteristics are usually utilized to identify those patients who need more critical care. However, the clinical evidence shows an unmet need to determine more accurate and optimal clinical biomarkers to triage patients under a condition like the COVID-19 crisis. Here we have presented a machine learning approach to find a group of clinical indicators from the blood tests of a set of COVID-19 patients that are predictive of poor prognosis and morbidity. Our approach consists of two interconnected schemes: Feature Selection and Prognosis Classification. The former is based on different Matrix Factorization (MF)-based methods, and the latter is performed using Random Forest algorithm. Our model reveals that Arterial Blood Gas (ABG) O_2_ Saturation and C-Reactive Protein (CRP) are the most important clinical biomarkers determining the poor prognosis in these patients. Our approach paves the path of building quantitative and optimized clinical management systems for COVID-19 and similar diseases.

## 1 Introduction

Upon the emergence of Coronavirus Disease 2019 (COVID-19), clinical-decision making to provide the best possible care to patients with this disease became an important issue. So far, more than 180 million cases of COVID-19 and 3,900,000 deaths due to it has been reported globally [1]. COVID-19 is a complex disease that affects different organ systems, and its clinical manifestations include a wide range of symptoms and signs [2, 3]. On the other hand, the clinical course of the disease is a complex phenomenon that can lead to death in some patients even when they do not have any comorbidity. Older age, accompanying chronic diseases, and imaging findings have been considered to worsen the prognosis, but they cannot predict the course of the disease and prognosis by themselves based on clinical observations in different patients’ populations [4, 5, 6, 7, 8, 9, 10]. The disease can cause a spectrum of acute or chronic complications that can perturb the trajectory of the disease progression and outcome [3, 4, 11].

In the era of systems medicine, Machine-Learning (ML) and Artificial Intelligence (AI) methods can be implemented to address clinical decision-making [12]. Big data analytics in systems medicine for this aim faces two major problems. The first issue is to preserve the geometric properties of the original data during the process of reducing the dimension of the data. While the original data are assumed to be sampled on a manifold of high-dimension and the function of reduction techniques is considered as mapping these data to a submanifold of a lower dimension in a way that the local geometry of the whole data is still included in the reduced data. The second problem is related to the noises and outliers in most clinical data which their negative impacts on the data analysis should be reduced or controlled effectively. There have been attempts to solve these problems in systems pharmacology using feature selection methods based on the Matrix Factorization (MF) [13].

To tackle the problem of missing geometric properties during reduction of the dimension and to soften the destructive influence of outliers and noises on data, many reduction techniques and tools have been offered so far. As a notable example, the category of subspace learning methods has received a significant attention due to the remarkable ability of such techniques to deal with the datasets of high-dimension, such as gene expression datasets. Subspace learning is a dimensionality reduction method that can produce a low-dimensional representation from the initial high-dimensional data. This method can be mixed with other techniques such as MF, manifold learning, and correlation analysis to perform both feature extraction and feature selection with excellent performance.

A successful example of the idea of subspace learning in unsupervised feature selection is Matrix Factorization Feature Selection (MFFS) method [14]. It carries out the feature selection in an iterative fashion using an algorithm based on non-negative matrix factorization and a subspace distance. Although MFFS was successful in bringing the matrix factorization from the world of feature extraction to the feature selection universe, it failed to consider the correlations among features. The latter leads to a feature subset with redundancy. Qi et al. [15] resolved the redundancy issue by introducing a new unsupervised feature selection method called the Regularized Matrix Factorization Feature Selection (RMFFS). RMFFS uses a regularization term, which is a combination of *L*_1_-norm and *L*_2_-norm, in optimizing the objective function of the matrix factorization. This approach results in a feature subset with low redundancy (i.e., linear independence) and a good representation of the original high-dimensional data. Alternatively, Wang et al. [16] introduced Maximum Projection and Minimum Redundancy (MPMR) as another unsupervised subspace learning method to reduce the redundancy in the selected features. MPMR formalizes the feature selection as a mapping from the feature space to the feature subspace using a projection matrix with the constraint of the minimum reconstruction error. Then, finding the projection matrix is reformulated as a matrix factorization problem that is solved using a greedy algorithm. To select low redundancy of the feature subset, a regularization term is added, which incorporates the Pearson correlation coefficient between features. None of the mentioned methods preserves the geometric structure of the features. To solve this issue, Shang et al. [17] presented the Subspace Learning-Based Graph Regularized Feature Selection (SGFS) method. SGFS incorporates graph regularization into subspace learning by constructing a feature map on the feature space. However, this method only preserves the geometry structure of the feature manifold. To preserve the geometric structures of both the feature and the data manifolds, Shang et al. [18] developed a new feature selection method called Sparse and Low-redundant Subspace learning-based Dual-graph Regularized Robust (SLSDR). SLSDR incorporates both feature and data graphs (dual graph) into subspace learning to select the feature subset that best preserves the geometric structures of both feature and data manifolds. The representativeness and low redundancy of the feature subset are guaranteed in SLSDR through the inner product regularization term. This is implemented in the feature selection matrix, which leads to sparse rows and the correlations between features being considered. Furthermore, SLSDR is robust against outliers, which is achieved by imposing *L*_2,1_-norm on the residual matrix of subspace learning.

This paper aims to revisit MFFS, MPMR, SGFS, RMFFS, and SLSDR, and to study their applications in two biomarkers and clinical data categories. First, to analyze ten gene expression datasets for the gene selection techniques. Second, to examine a COVID-19 clinical dataset by extracting its predictive features and to present a model to discover clinical signatures of poor prognosis in COVID-19. The main aim to use the techniques mentioned above is their significant performance in handling feature selection problems. To be specific, the feature selection mechanism developed for MFFS has been demonstrated to be highly efficient and productive, so that a broad category of techniques has been founded on the MFFS framework. MPMR, SGFS, RMFFS and SLSDR fall in this category and they improve the performance of MFFS from different perspectives using different tools. These powerful tools to develop feature selection methods include subspace learning, non-negative matrix factorization, manifold learning, and correlation analysis. A chronological and detailed illustration of the framework for the methods MFFS MPMR, SGFS, RMFFS, and SLSDR is shown in Figure 1.

**Figure 1:**
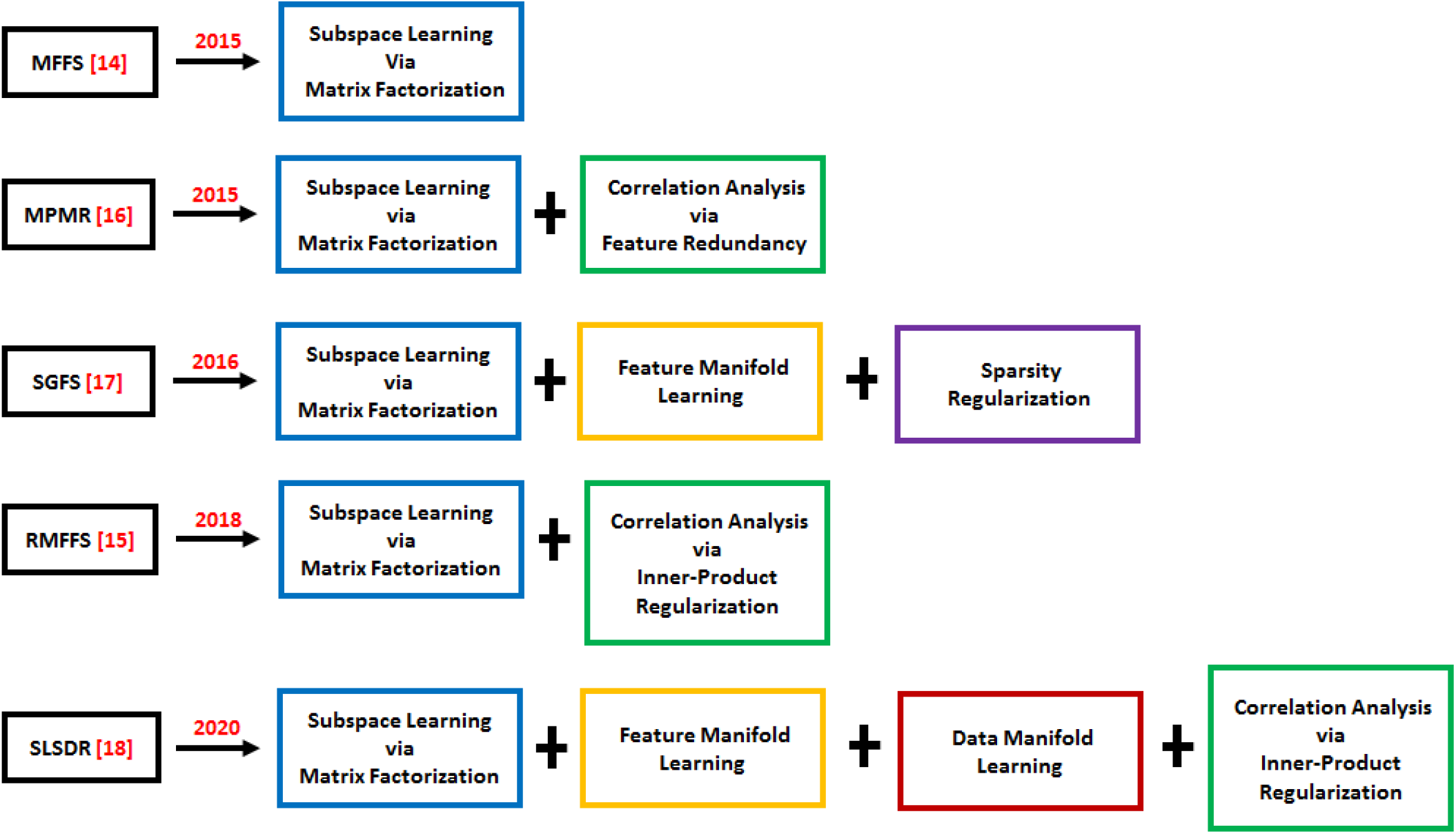
Chronological and detailed illustration of the basic framework for the methods MFFS MPMR, SGFS, RMFFS and SLSDR.

The organization of the subsequent sections is as follows. Section 2 provides descriptions regarding the taxonomy and the different insights utilized in this study. In Section 3, a detailed description of the mechanisms of MFFS, MPMR, SGFS, RMFFS and SLSDR is provided and the benefits and drawbacks of each of these techniques are studied. In Sections 4 and 5, several experiments are conducted on a set of benchmark gene expression datasets and a COVID-19 clinical dataset. Moreover, a comprehensive analysis of the obtained results is carried out. Finally, Section 6 concludes the paper.

## 2 Taxonomy

This section presents an explication of the taxonomy of our work and describes the perspectives on the feature selection techniques.

### 2.1 Dimensionality Reduction

Massive datasets that are composed of features of high-dimension and include relatively low number of patterns pose critical challenges to machine learning techniques [19]. Dimensionality reduction is an important problem in machine learning applications in high-dimensional datasets. Specifically, when there are a number of redundant or irrelevant features in an initial feature set [20], reducing the dimensionality of high-dimensional data is often necessary for two main reasons. Firstly, it helps reduce the computational complexity and memory usage. Secondly, high-dimensional datasets have some redundant and noisy features that can negatively impact the performance of machine learning models. Therefore, selecting a subset of relevant features can reduce the computational cost and lead to models that generalize better.

Two main categories of dimensionality reduction techniques have been introduced [21, 22]: Feature Selection and Feature Extraction. The former includes methods that select a subset of relevant features that represent the original data, whereas the latter is focused on the transformation or projection of the original data that approximate all features. Feature extraction incorporates the original features to make new features with lower dimension that contain all or at least the major pieces of the information included in the initial features. Feature selection aims to select a subset of the original features by removing the irrelevant and redundant features. As a result, while the feature selection methods only need collecting selected features, feature extractions require all features for dimensionality reduction.

Feature selection methods can be supervised, unsupervised and semi-supervised, depending on whether the dataset has label information or not [21, 22, 23, 24]. Supervised learning methods select relevant features by evaluating the correlation between the features and their corresponding label. Fisher score [25], Hilbert Schmidt Independence Criterion (HSIC) [21], trace ratio criterion, and mutual information [26] are among the most common supervised feature selection methods. Discriminative information inherent in the label data facilitates selecting discriminant features from the original ones. When the dataset is partially labeled, semi-supervised methods are used to handle feature selection. These methods evaluate the feature relevance based on both the discriminative information of labels and the information embedded in the local structure of the whole dataset. However, there are situations where obtaining sufficient labeled data is hard. To deal with unlabeled datasets, a variety of unsupervised feature selection techniques such as MF have been developed [14, 27]. In contrast to two previous methods, the unsupervised techniques do not have access to the discriminative information inherent in the labeled data. In this case, the feature selection from unlabeled data is challenging. We introduce various unsupervised feature selection methods applied to analyze some unlabeled datasets.

Based on the searching algorithm for selecting relevant features, the feature selection methods can be categorized into four groups: Filter, Wrapper, Embedded and Hybrid methods [21, 28]. Filter methods use the inherent properties of the data to evaluate the feature importance and to assess the appropriateness of features without using any machine learning algorithm. For this aim, filter methods use some ranking metrics such as Laplacian score, feature similarity, and trace ratio. On the other hand, wrapper methods applies specific learning algorithms like classification or clustering to select the most relevant feature subset that results in better performance of the utilized learning algorithm. In this type of feature selection, a search technique is employed to find the best feature subset. At each iteration, this technique produces a number of candidate feature subsets and evaluates the usefulness of each generated subset using a classification or a clustering algorithm. The subset assessed as the most efficient is considered as the final feature set [23, 29]. The advantages of filter methods compared to wrapper methods are higher scalability and lower computational cost [30, 31, 32]. Embedded methods carry out the feature selection during the model construction process. Since embedded methods often consider various characteristics of data such as the local manifold structure, they provide better performance in feature selection compared to the filter and wrapper methods [33]. Hybrid models are constructed by the incorporation of the filter-based techniques into wrapper-based methods aiming to use the advantages of both models [34, 35].

### 2.2 Matrix Factorization

Matrix factorization (MF) is a well-known mathematical scheme that has recently been applied to a range of complex problems in the numerical linear algebra, machine learning and computational biology. Some notable examples include Eigendecomposition of a matrix [36], Data Compression [37], Recommender Systems [38], Spectral Clustering [24], and gene expression analysis [39]. Some of the MF-based techniques that have been used widely are Singular Value Decomposition (SVD) [40], Principle Component Analysis (PCA) [41], and Probabilistic Matrix Factorization (PMF) [42]. The focus of the latest lines of ML research is to utlize those MF methods that are particularly capable of finding patterns appropriate for data interpretability using some essential properties of data. Especifically, non-negative matrix factorization (NMF) [27] is used to analyze matrices of data with non-negative entries. These matrices and decomposing them are common in the context of image and text datasets analysis.

Let **X** ∈ ℝ^*n*×*d*^ be a non-negative data matrix. The NMF technique, seeks a parts-based representation of **X** in terms of two low-rank non-negative matrices. The general formulation of the NMF can be expressed as **X** ≈ **PQ** in which the two non-negative matrix factors **P** ∈ ℝ^*n*×*k*^ and **Q** ∈ ℝ^*k*×*d*^, called the basis and the coefficient matrix respectively, represent **X**. Furthermore, it is recently proven that from a theoretical perspective, NMF is closely connected to the k-means clustering algorithm. For this reason, NMF is noted as one of the best unsupervised learning methods in identifying the latent subspace of data and is particularly appropriate in data clustering [43]. Over the past decades, many techniques have been founded on the mechanism of NMF including the conventional NMF [27], the convex NMF (CNMF) [44], the orthogonal NMF (ONMF) [45] and the semi-NMF (SNMF) [44]. Accordingly, the constraint of nonnegativity imposed on the data and the basis matrix in NMF is relaxed in the framework of SNMF, the elements of the basis in CNMF are assumed to have a representation in the form of a convex combination of the input data vectors, and the factor matrices in ONMF are constrained by the orthogonality condition to guarantee the interpretation of the clustering.

It should be pointed out that the NMF-based methods achieve strong and economic performance which is a leading factor in the widespread use of these techniques in many research fields, and particularly in computational biology [46, 47, 48]. A large number of studies in physiology and neuropsychology have presented evidence to propose that representing a non-negative matrix corresponding to a dataset by parts-based factors should be a proper approach to analyze the recognition system of the human brain [43]. NMF-based methods can be used for mathematical models of diseases to find and fine-tune the parameters of the models using experimental or clinical data [49].

### 2.3 Subspace Learning

Subspace learning is another way of dimensionality reduction that assumes the input data lies on some intrinsic lower dimensional space to which the original features are mapped. Specifically, subspace learning can be considered as a powerful tool to represent a space of a higher dimension by a subspace of a lower dimension using a learning technique. Principal Component Analysis (PCA) [41, 50], Linear Discriminant Analysis (LDA) [51, 52], Locality Preserving Projection (LPP) [53], and Neighborhood Preserving Embedding (NPE) [54] are among the most common subspace learning methods. All of these techniques can be considered as a variant of manifold learning that linearly projects the input data into a subspace embedded in the ambient space. The linear mapping of these methods makes them faster than non-linear variants of manifold learning such as Locally Linear Embedding (LLE) [55]. Unlike PCA that preserves the global euclidean structure of the input data, the neighborhood structure of each data point is preserved in NPE. This feature of NPE is similar to LPP, although their objective functions are different from each other.

The reduction techniques described in this section are not proper to deal with feature selection tasks since their mechanisms are developed only to handle feature extraction problems. It seems likely that the representation provided by the features in the subspace of a lower dimension would not be well interpretable. To overcome this limitation, MF, NMF and other subspace learning conceptions have been merged to develop a broad category of novel and effective feature selection techniques. Some significant methods introduced recently in this category include SL-IGO [56], MFFS [14], MPMR [16], NSSLFS [57], SGFS [17], GLOPSL [58], LSS-FS [59], RMFFS [15], SFS-BMF [60], RNE [61], SLSDR [18], and SLASR [62].

### 2.4 Manifold Learning

Manifold learning uncovers low-dimensional manifolds (constraint surfaces) that are embedded in the high-dimensional space of the input data in an unsupervised manner. Thus, a manifold learning method leads to a non-linear dimensionality reduction in the input data with high dimensionality such as medical images or high-spectral remote sensing data. The resulting low-dimensional embedding best preserves the manifold structure of the original data. To verify this condition, some statistical measures such as variance or reconstruction error is usually used. Depending on the type of the statistical measure being utilized, various manifold learning methods have been developed. Some early examples include Isometric Feature Mapping (Isomap) [63], Locally Linear Embedding (LLE) [55], Laplacian Eigenmaps [64], Laplacian Score [65], and Multi-Cluster Feature Selection (MCFS) [66]. These methods take into account the manifold structure of the data through either their explicit formulation and/or some sort of regularization. All of these methods only consider the manifold structure of the samples.

Dual-manifold learning approaches have been also used recently. The focus of dual-manifold learning approaches is to use the samples and features duality connection to exploit the manifold structures of both samples and features of the original data. The idea behind the manifold learning and dual-manifold learning approaches is to include the samples and/or features geometric structure in the reduced data obtained as a result of the feature selection technique used. For the samples and/or features, an affinity graph is constructed which models their local geometric properties during the process of selecting features. Recently, many new and efficient feature selection methods have been founded on manifold learning and dual-manifold learning including SGFS [17], GLOPSL [58], RGNMF [67], DSNMF [68], DSRMR [69], DGRCFR [70], DGSPSFS [71], DMvNMF [72], LRLMR [73], SLSDR [18], MRC-DNN [74], RML-RBF-DM [75], and EGCFS [76].

### 2.5 Correlation Analysis

As a statistical tool, the correlation analysis examines two variables to determine the possible relationships between them. These variables may be both dependent or both independent. It is also possible that only one variable is dependent and the other is independent [77]. This analysis evaluates the variables connection making use of a criterion called the correlation coefficient. A positive or a negative value of this criterion indicates that a positive or a negative correlation exists between the two corresponding variables, respectively. Moreover, a greater or a smaller value of the correlation coefficient shows that a stronger or a weaker correlation exists between the corresponding variables, respectively [78, 79].

The information that the correlation of a feature set reveals has played a pivotal role in the framework of newly established feature selection techniques. It is demonstrated that this issue reflects new aspects of the original data which enhances the learning performance [21, 22]. The correlation-based feature selection approaches aim to explore the level of correlation among features to decide which features are connected and should be eliminated. The learning process is guided to minimize the correlation among the original data. The correlation corresponding to a set of features can not only determine the relevant features, but detect the feature redundancy [80]. In the selection process of the supervised techniques, the connection that every input feature may have with the target variable is investigated. For this purpose, some tools from statistics are applied including the mutual information [26], Pearson correlation coefficient [79], and Fisher score [25]. Using these notions, the selection of the input features is guided so that the most correlated features to the target are chosen. Regarding the unsupervised techniques, the selection process can be rested on two major frameworks that aim to compute the feature redundancy for an especial subset. The first framework applies some tools developed in information theory or statistics to calculate the level of pair-wise correlation, similarity and dependence of features. Some notable techniques that fit this framework can be found in [81, 82, 83, 84, 85, 86, 87]. The other framework uses a notion that is able to identify the features redundancy to calculate the features connections. TAn objective function is formed to assess the features in a jointly manner. The optimization task is regularized subject to a sparsity constraint. A number of important methods that fall into this category can be found in [15, 16, 18, 87, 88, 89, 90, 91]. Table 1 summarizes the different categories studied in this section.

**Table 1:** A summary of the taxonomy and references related to the feature selection methods revisited in this paper.

| Category | Description | References |
| --- | --- | --- |
| **Matrix Factorization**<br>**or**<br>**(Non-Negative Matrix**<br>**Factorization)** | To decompose a given (non-negative) matrix into the product of two low-rank (non-negative) matrices | [24, 27, 36, 37, 38, 39, 40, 41, 42, 43, 44, 45, 46, 47, 48] |
| **Subspace Learning** | To learn a low dimensional representation of a high-dimensional space | [13, 14, 15, 16, 17, 18, 41, 50, 51, 53, 54, 55, 56, 57, 58, 59, 60, 61, 62, 92] |
| **Manifold Learning** | To uncover low-dimensional manifolds that are embedded in the high-dimensional space of the input data | [17, 55, 58, 63, 64, 65, 66, 67, 73, 74, 76, 93] |
| **Dual-Manifold Learning** | To take into account the duality between samples and features and exploit the manifold structures of both samples and features of the original data | [18, 68, 69, 70, 71, 72, 75, 94, 95, 96, 97, 98] |
| **Correlation Analysis** | To identify the relationships between two variables | [15, 16, 18, 25, 26, 78, 79, 81, 82, 83, 84, 85, 86, 87, 88, 89, 90, 91] |

## 3 Background and Methods

In this section, five feature selection methods founded on a set of different concepts including the matrix factorization technique, the feature redundancy, the feature correlation, the data manifold and the feature manifold are described. These methods are compared and theoretical insights on their applications are described.

### 3.1 Notations

The data matrix **X** is described as **X** = [**x**_1_; **x**_2_; … ; **x**_*n*_] = [**f**^1^, **f**^2^, …, **f**^*d*^] ∈ ℝ^*n*×*d*^ in which *n* and *d* denote the number of samples and that of features, respectively. The notation **I**_*k*_ indicates the identity matrix of size *k*, and **1**_*k*_ denotes a square matrix of size *k* whose entries are all one. For any matrix **Z** ∈ ℝ^*m*×*n*^, the transpose of **Z** is denoted by Tr(**Z**). Moreover, the Frobenius norm and the *L*_2,1_-norm of **Z** are defined as 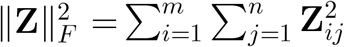, and 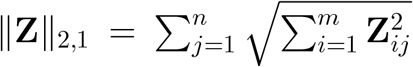, respectively. The Euclidean inner product of the two matrices **S, Z** ∈ ℝ^*m*×*n*^ is also presented by 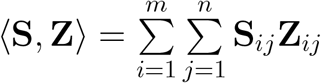.

### 3.2 MFFS

Two well-known techniques, MF and NMF, have been proven to be able to handle the clustering task and large-scale data processing in an efficient and productive manner. The mechanism of NMF, which is founded on the framework of MF, is to decompose a nonnegative matrix into two nonnegative matrices. Specifically, the nonnegativity condition effectively constrains MF so that only a part of the data representation is applied to handle the learning process. Recently, many innovative modifications, rested on MF and NMF, have been incorporated into the feature selection framework. Wang et al. proposed “Matrix Factorization based Feature Selection” (MFFS) as a new selection method by applying the matrix factorization to a subspace distance measure [14]. The MFFS technique helps to solving the minimization problem given as follows:

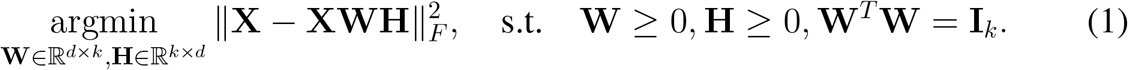

The feature selection mechanism in MFFS is in fact the MF process in which the corresponding optimization framework is constrained by orthogonality condition. Since solving Problem (1) is a challenging task, the term **W**^*T*^ **W** = **I**_*k*_ is constrained by including a penalty term in the problem presented by Eq. (1). Therefore, Problem (1) is modified as follows:

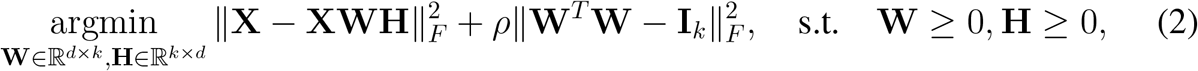

in which *ρ* denotes the balancing coefficient for the penalty term. A major difficulty in dealing with Problem 2 is that while the objective function given in Eq. (2) separately satisfies the convexity condition with respect to **W** or **H** when **H** or **W** is fixed respectively, it does not fulfill that condition for both **W** and **H** simultaneously. In order to solve Problem (2), the Lagrange multiplier method is incorporated into the optimization framework used. Particularly, all the variables are taken to be constant except for the one that is optimized. The optimization process is established on an iterative algorithm for which there is a convergence criterion to determine when the algorithm should stop. Algorithm 1 summarizes the MFFS framework.

#### Algorithm 1 The MFFS method.

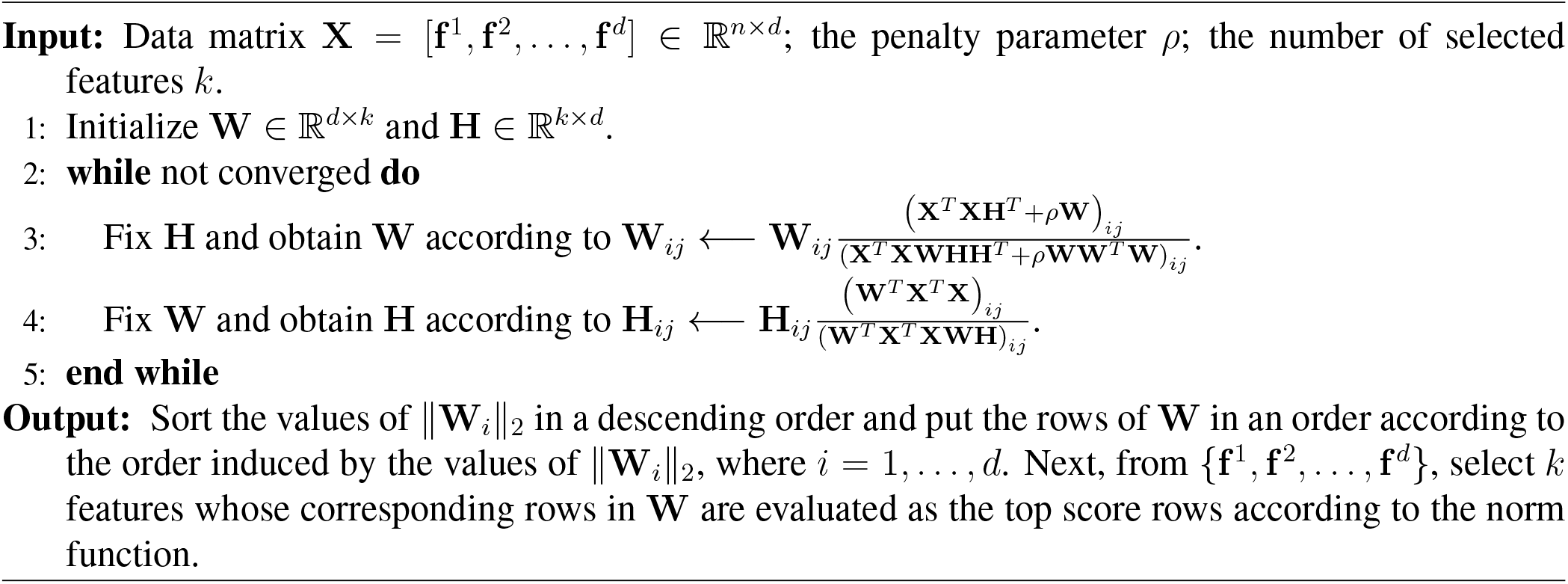

### 3.3 MPMR

A major traditional function of feature selection is to remove those features that are irrelevant in a given dataset. However, handling mining processes by feature selection techniques is a complex problem when the data are of a high-dimension. Such datasets can be noisy and include a large number of features that are redundant or irrelevant. To overcome this problem in mining performance, strategies to minimize the data redundancy can be incorporated into feature selection for the high dimensionality case. Wang et al. [16] introduced “feature selection based on Maximum Projection and Minimum Redundancy” (MPMR) as a new and efficient selection technique rested on evaluation of features redundancy. This technique detects how much a given feature is relevant to a subset of features. MPMR modifies the objective function of MFFS presented by Eq. (1) in Section 3.2 to develop a feature selection framework based on minimization of the redundancy between the selected features. The optimization problem of MPMR is:

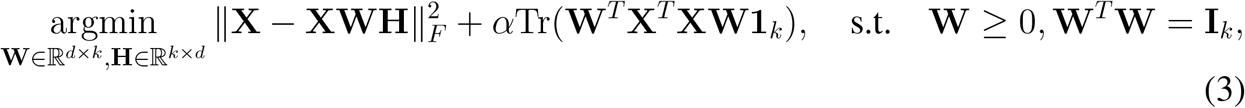

where *α* provides an appropriate balance between the degrees of approximation and redundancy, *k* is the number of the selected features, and the redundancy rate for the selected features is represented by the term Tr(**W**^*T*^ **X**^*T*^ **XW1**_*k*_) in which **1**_*k*_ denotes a square matrix of size *k* whose entries are all one. An interesting issue is that Problem 3 is free from the constraint **H** ≥ 0 imposed on Problem 2. The objective function given in Eq. (3) is handled in an iterative manner through two steps. First, **H** is taken to be constant while **W** is updated to be optimized. Next, the optimal **W** is applied to optimize **H**. In Algorithm 2, the framework for minimizing the objective function of MPMR is presented.

### 3.4 SGFS

An important property common among many datasets of high-dimension is being locally structured. It is demonstrated that the local geometry of such data has profound and constructive impacts on enhancement of the learning techniques performance. To work with high-dimensional data, one needs to reduce the dimension while preserving the local structure. To this aim, high dimensional data are mapped into a subspace of lower dimension of the original data space so that the projected data still contain the local properties of the original data. A majority of feature selection techniques use the geometry preservation task by the Laplacian graph. As an example, the “Subspace Learning-Based Graph Regularized Feature Selection” (SGFS) technique newly proposed by Shang et al. [17] applies the feature manifold conception to the MFFS framework discussed in Section 3.2. It is remarkable that representing the feature manifold by a feature graph, SGFS effectively addresses the problem of missing local geometrical structure in the frameworks of MFFS and MPMR.

#### Algorithm 2 The MPMR method.

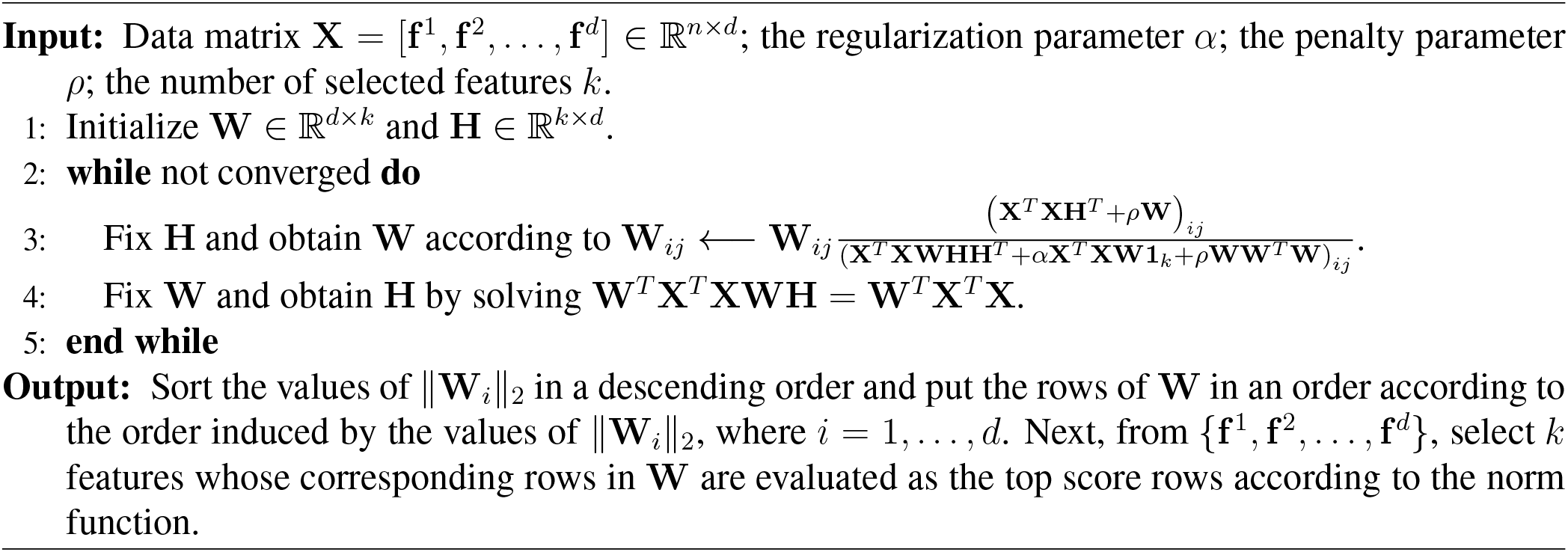

#### Feature Graph

Suppose that *G*^*F*^ is a feature graph whose set of vertices corresponds to the original features set {**f**^1^, **f**^2^, …, **f**^*d*^}. Moreover, let for 1 ≤ *i, j* ≤ *d*, the feature similarity values, denoted by 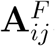, are calculated as:

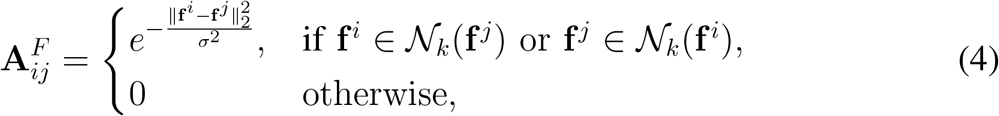

where *σ*, known as the Gaussian parameter, is a neighborhood size controlling parameter, and 𝒩_*k*_(**f**^*i*^) is a set called the *k*-nearest neighbor of **f**^*i*^. Each 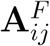 is considered as a weight for an edge *G*^*F*^ and indicates how similar two features **f**^*i*^ and **f**^*j*^ are. In particular, the larger 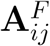 is, the more similar the features **f**^*i*^ and **f**^*j*^ will be. In the next step, for the feature manifold, the graph Laplacian matrix **L**^*F*^ is constructed making use of the similarity matrix 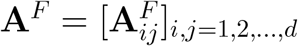. The Laplacian matrix is defined as **L**^*F*^ = **D**^*F*^ − **A**^*F*^ in which **D**^*F*^ is a diagonal matrix whose diagonal entries are 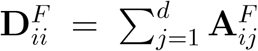 for 1 ≤ *i* ≤ *d*.

It should be noted that the matrix **H** given in Eq. (1) provides a practical criterion to assess the features similarity so that a high level of similarity between the features **f**^*i*^ and **f**^*j*^ implies more similarity between the columns **h**^*i*^ and **h**^*j*^ which represent **f**^*i*^ and **f**^*j*^, respectively [17]. Incorporation of a feature graph into the framework of feature selection that uses the mentioned fact to guide the construction process of **H** can be formulated as:

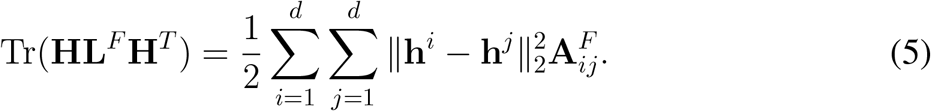

The optimization problem for SGFS is obtained by introducing the feature graph term presented in Eq. (5) to Problem (1) as:

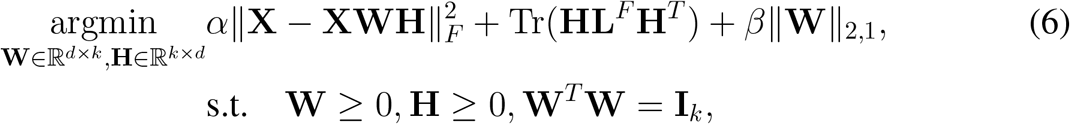

where the nonnegative parameters *α* and *β* provide a trade-off between the terms, and the sparsity of the matrix **W** is ensured by placing the *L*_2,1_-norm as a constraint on **W**. To deal with this constraint in the calculations, the formula ‖**W**‖_2,1_ = Tr(**W**^*T*^ **PW**) can be applied in which the diagonal matrix **P** is defined in terms of its diagonal entries 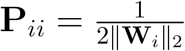, where 1 ≤ *i* ≤ *d*. Based on Algorithm 3, Problem (6) is solved separately for each variable assuming that the rest of variables are taken to be fixed.

##### Algorithm 3 The SGFS method.

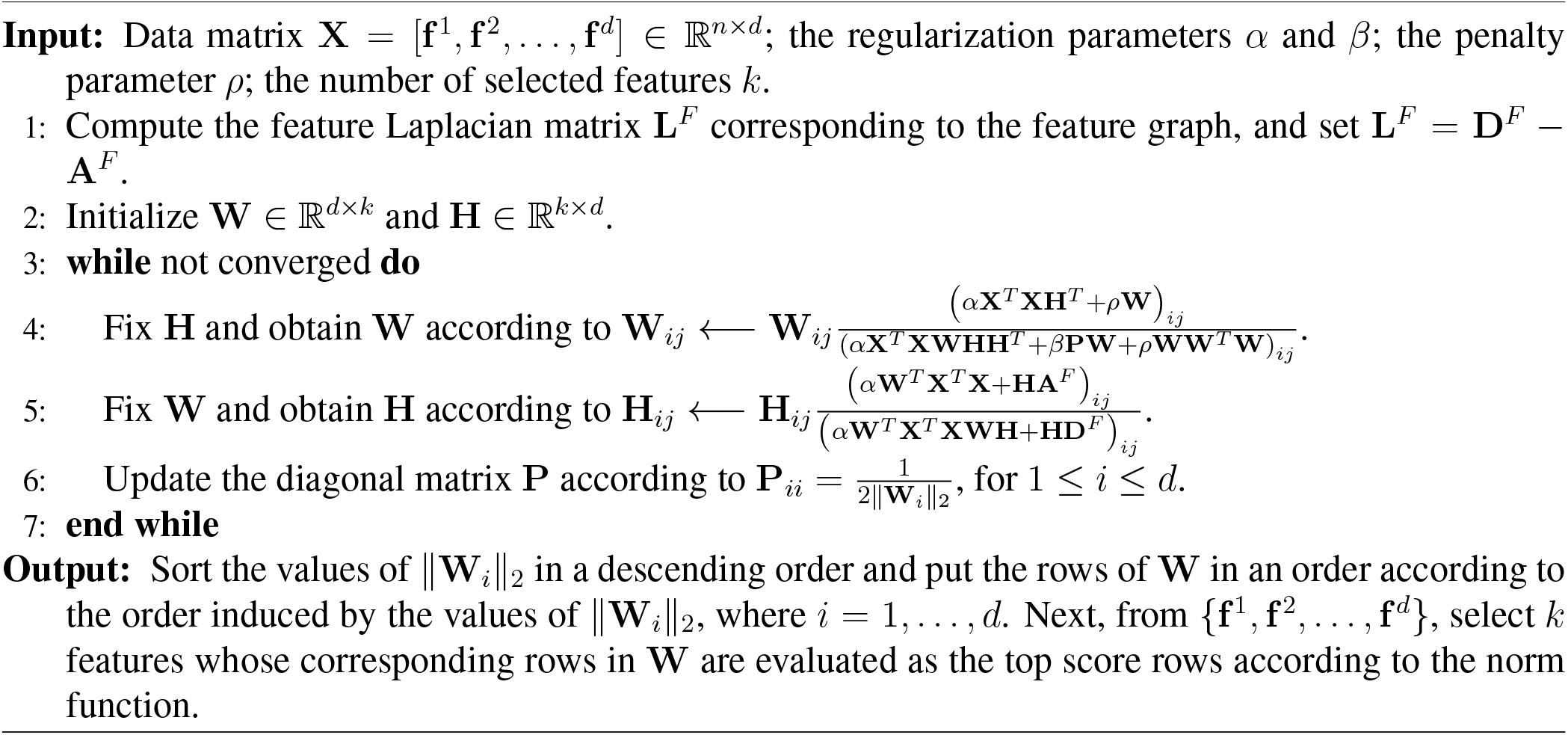

### 3.5 RMFFS

As discussed before, the feature selection frameworks for MFFS, MPMR and SGFS were developed based on imposing an orthogonality constraint on the feature weight matrix **W**. In practice, this constraint is hardly fulfilled since orthogonality is normally too strict. Furthermore, there are more drawbacks regarding the framework of the mentioned techniques. A major downside of MFFS and SGFS is that the correlations among the features are disregarded in MFFS and SGFS frameworks and this issue can negatively affect the process of selecting discriminative features. Another drawback of SGFS is the ignorance of redundancy caused by the *L*_2,1_-norm which constrains the feature weight matrix to regularized it so that the feature selection is performed in a more efficient way. Several nformative features that are redundant may be disregarded by SGFS since the measurement of redundancy is neglected by the *L*_2,1_-norm.

These problems are solved in the framework of “Unsupervised Feature Selection by Regularized Matrix Factorization” (RMFFS) [15]. This method uses the non-negative matrix factorization structure used by MFFS, MPMR and SGFS to propose a novel feature selection technique in which an inner product regularization term associated with the feature weight matrix is exerted into the objective function of RMFFS. The major contribution of RMFFS is that the sparsity of the feature weight matrix and the low redundancy among the selected features is guaranteed at the same time. The optimization problem for RMFFS is constructed as:

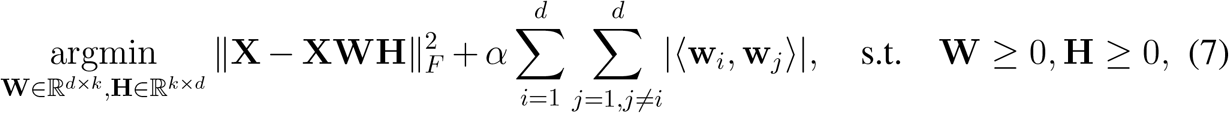

in which *α* is a trade-off parameter. By a simple calculation, Problem (7) is expressed as:

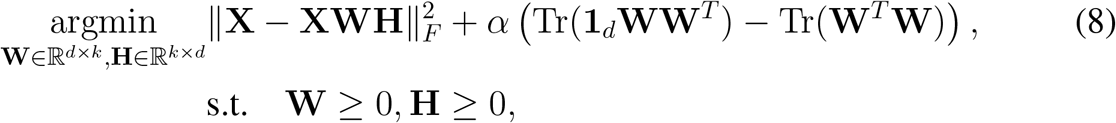

which is more straightforward to be calculated. In Eq. (8), **1**_*d*_ represents a square matrix of size *d* with one as its entries everywhere.

Problem (8) can be solved by updating a single variable until convergence takes place while the rest of the variables are taken to be fixed. convergence is repeated. Algorithm 4 summarizes the RMMFS framework.

### 3.6 SLSDR

The geometric information locally embedded in both the data and feature manifolds can play important roles in the improvement of dimensionality reduction performance [99, 100]. Despite of this fact, there are only a handful of methods that apply both feature and data geometries into the feature selection. Sparse and Low-redundant Subspace Learning-based Dual-graph Regularized Robust Feature Selection, “SLSDR” [18], was proposed by making an extension on the MFFS and SGFS techniques [14, 17] so that the data and feature manifolds, represented by dual-graph regularization terms, were included in the formulation of SLSDR at the same time. This way, SLSDR selects those features that can best represent the geometric aspects of the whole data.

#### Algorithm 4 The RMFFS method.

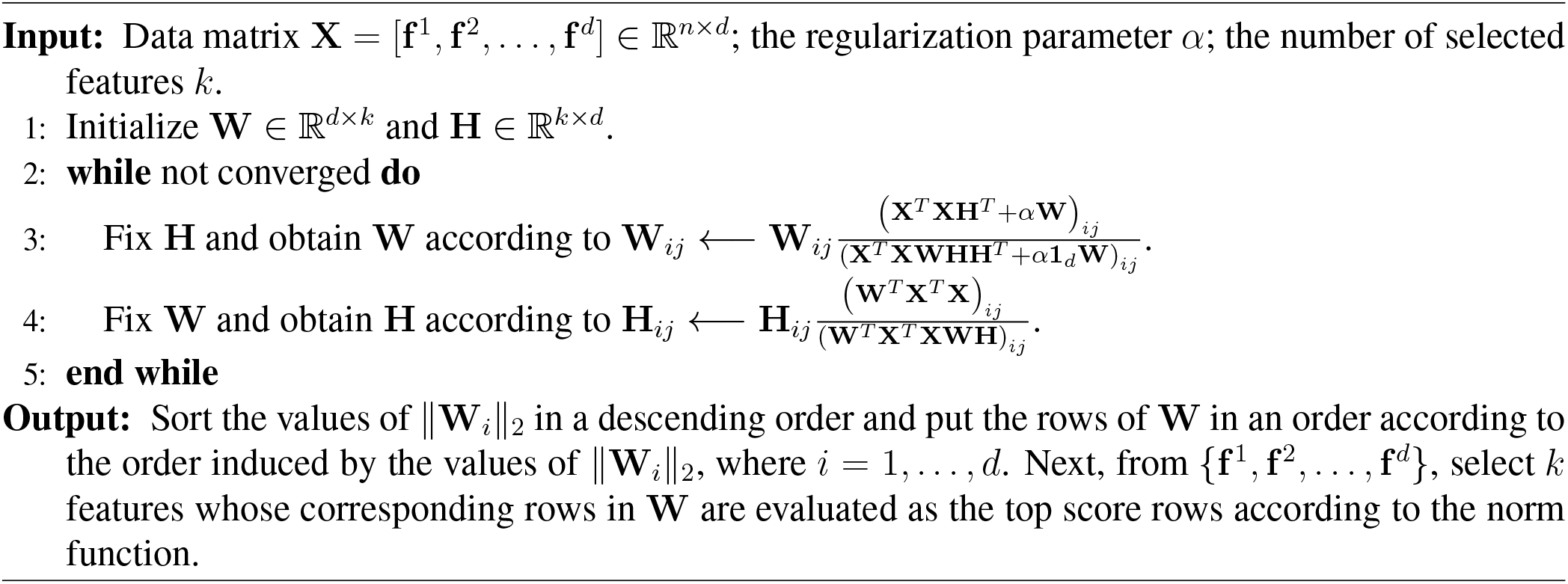

It is apparent that in SLSDR, both graphs of samples and features are built to detect the geometry of samples and features, respectively. SLSDR utilizes the same feature graph construction strategy as the one discussed for SGFS in Subsection 3.4. The data graph construction strategy used by SLSDR is described in details as follows.

#### Data Graph

Similar to the MFFS feature selection framework, the matrix of feature selection, **W** ∈ ℝ^*d*×*k*^, is employed in SLSDR in order to include the local geometric information of data in the selection algorithm. To be more specific, assume that the graph of the *k*-nearest neighbor, applied to create the manifold of data in an efficient manner, is represented by 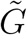 in which the set of vertices is identified as {**x**_1_, **x**_2_, …, **x**_*n*_} so that each vertex of the graph corresponds to a sample *x*_*i*_, for 1 ≤ *i* ≤ *n*. In the same way as the feature graph was constructed, the data similarity weights can be introduced. In particular, for 1 ≤ *i, j* ≤ *n*, these weights are associated to each edge, which links two vertices **x**_*i*_ and **x**_*j*_, to compute the similarity between **x**_*i*_ and **x**_*j*_ as:

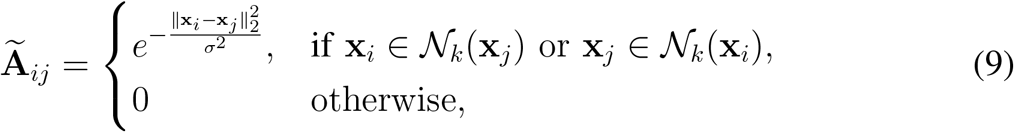

where *σ* denotes the Gaussian parameter which determines the neighborhoods length, and 𝒩_*k*_(**x**_*i*_) is the *k*-nearest neighbor set of **x**_*i*_. It should be pointed out that **Ã**_*ij*_ is utilized to compute the similarity between **x**_*i*_ and **x**_*j*_. In other words, a higher value of *Ã*_*ij*_ implies a greater similarity between **x**_*i*_ and **x**_*j*_. Then, for the data manifold, the Laplacian matrix, 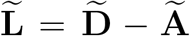, is calculated in which 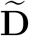 is diagonal with 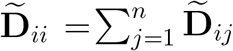 as its diagonal entries, and **Ã** = [**Ã**_*ij*_].

As the data manifold assumption [18] states, for two samples **x**_*i*_ and **x**_*j*_ whose similarity is high, the linear mappings **x**_*i*_**W** and **x**_*j*_**W** correspond to **x**_*i*_ and **x**_*j*_, respectively, share strong similarity. The data graph in terms of the matrix **W** is formulated by:

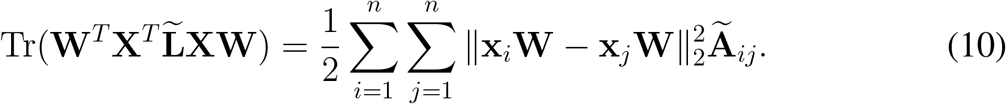

Furthermore, motivated by the benefits that the regularization technique provided for RMFFS, a term for regularizing the selection matrix is added to the feature selection model of SLSDR in the form an inner product. This new term aims to determine the most representative features whose redundancy is low, and as a consequence, both the rows sparsity and the correlations between the features are promisingly included in the selection process. In summary, the objective function of SLSDR is given by:

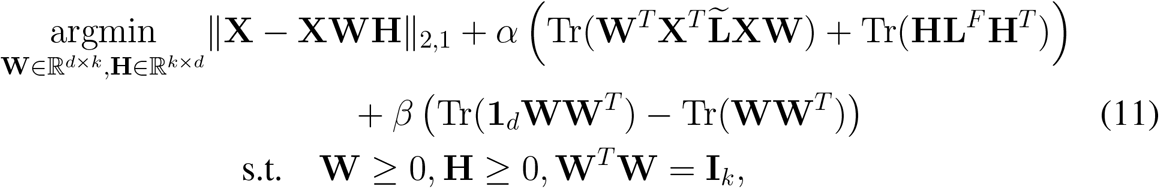

in which *α* and *β* are two balance parameters, **1**_*d*_ indicates a square matrix of dimension *d* with one as its entries everywhere, and the *L*_2,1_-norm is utilized to guarantee the intensity of the subspace learning residual matrix against outliers. Moreover, **L**^*F*^ denotes the Laplacian matrix associated with the manifold of features described in Subsection 3.4 by the feature graph.

Problem (11) is handled using the framework presented by Algorithm 5. As discussed in the former subsections, the optimization process is rested on updating only one variable while the other variables are fixed until a convergence criterion is satisfied.

## 4 Experimental results

In this section, we present the experimental results to evaluate the efficiency and efficacy of five different feature selection algorithms which include MFFS, MPMR, SGFS, RMFFS, and SLSDR. These algorithms have been applied to ten publicly available gene expression datasets that are summarized in Table 2. Some of these results are taken from our previous work [13].

**Table 2:**
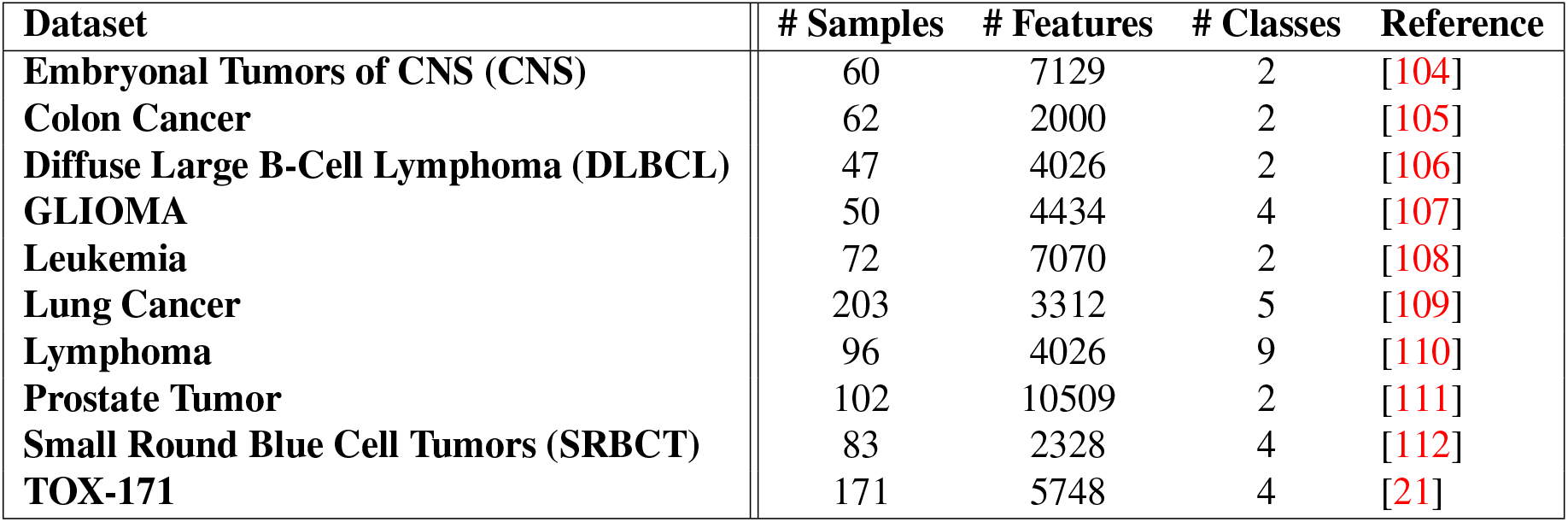
Details of ten gene expression datasets used in the experiments.

| Dataset | # Samples | # Features | # Classes | Reference |
| --- | --- | --- | --- | --- |
| Embryonal Tumors of CNS (CNS) | 60 | 7129 | 2 | [104] |
| Colon Cancer | 62 | 2000 | 2 | [105] |
| Diffuse Large B-Cell Lymphoma (DLBCL) | 47 | 4026 | 2 | [106] |
| GLIOMA | 50 | 4434 | 4 | [107] |
| Leukemia | 72 | 7070 | 2 | [108] |
| Lung Cancer | 203 | 3312 | 5 | [109] |
| Lymphoma | 96 | 4026 | 9 | [110] |
| Prostate Tumor | 102 | 10509 | 2 | [111] |
| Small Round Blue Cell Tumors (SRBCT) | 83 | 2328 | 4 | [112] |
| TOX-171 | 171 | 5748 | 4 | [21] |

### Algorithm 5 The SLSDR method.

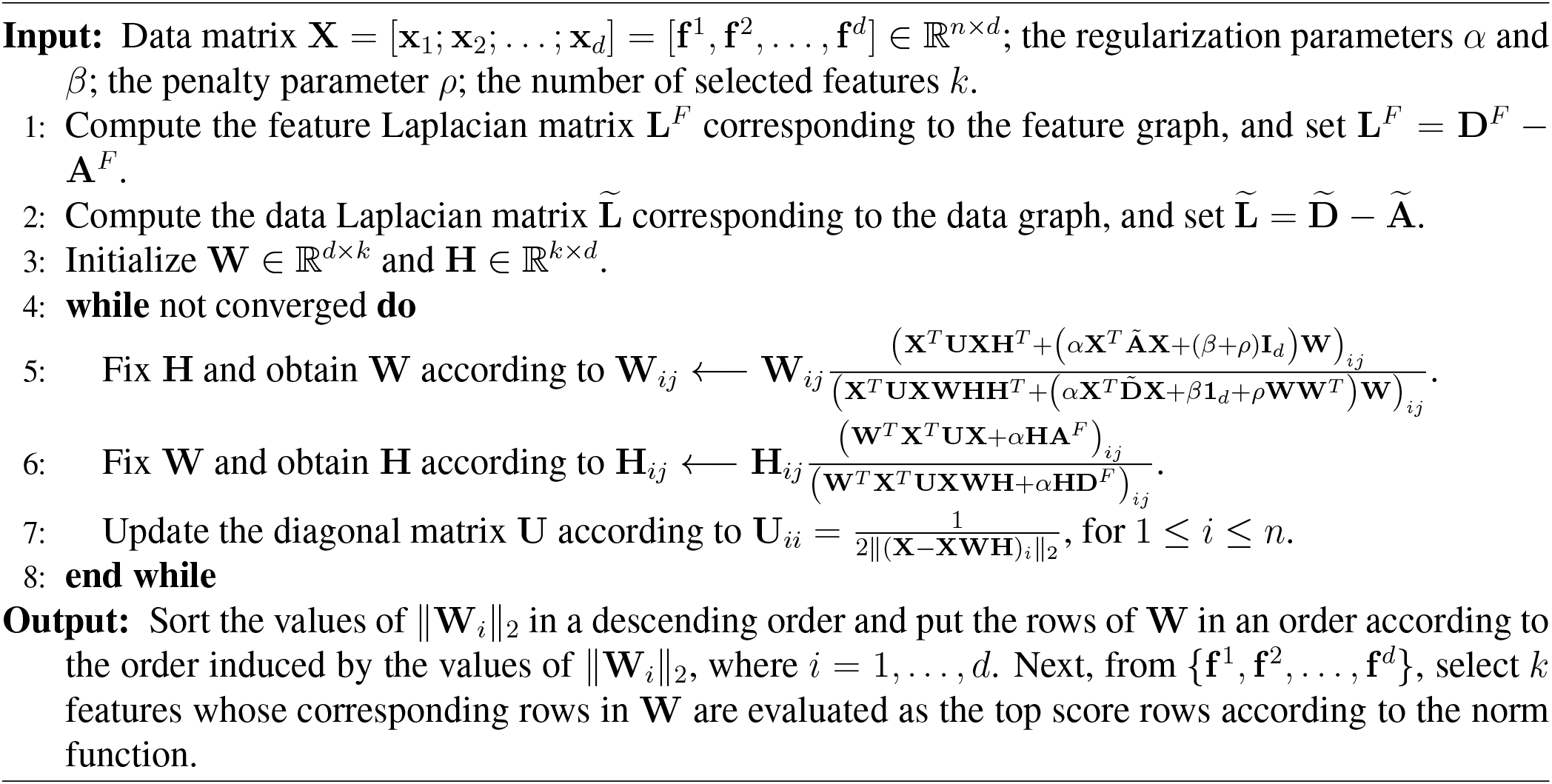

### 4.1 Description of Datasets

Below, the datasets that we have used in our experiments are briefly described. It should also be noted that these datasets are accessible in the Repositories [101, 102, 103].

1. **Embryonal Tumors of CNS (CNS)** dataset [104] contains 60 records for the outcome prediction of patients dealing with central nervous system embryonal tumor and includes 7129 gene expressions as features for each patient. 21 of the records correspond to survivors and the rest are cases of morbidity.
2. **Colon Cancer** gene expression dataset [105] includes 62 biopsies taken from patients with colon cancer. Each record in the dataset, which has 2000 gene expressions as features, is labeled as either “negative” or “positive”. The former corresponds to biopsies taken from tumors, while the latter indicates normal biopsies taken from the normal tumor tissue.
3. **Diffuse Large B-Cell Lymphoma (DLBCL)** [106] is a gene expression dataset, including 4026 gene expressions as features, for distinct types of DLBCL. The dataset is made up of 47 samples out of which 24 are from “germinal center B-like” group while the rest are “activated B-like” group.
4. **GLIOMA** [107] is a dataset of four classes labels and 50 samples with 4433 gene expressions as features. The four class labels are: cancer glioblastomas (CG), non-cancer glioblastomas (NG), cancer oligodendrogliomas (CO), and non-cancer oligodendrogliomas (NO). The number of samples corresponding to these class labels are 14, 14, 7, and 15, respectively. Thus, this dataset is relatively balanced with respect to the number of data points for each class.
5. **Leukemia** dataset [108] is collected from 72 bone marrow samples of 47 Acute Lymphoblastic Leukemia (ALL) patients and 25 Acute myeloid Leukemia (AML) patients. It is a binary dataset in which each sample has 7070 gene expressions as features.
6. **Lung Cancer** [109] is also a gene expression, multi-class dataset of 203 samples, 3312 gene expressions as features, and five distinct class labels. The classes labels are adenocarcinomas, squamous cell lung carcinomas, pulmonary carcinoids, small-cell lung carcinomas and normal lung, with 139, 21, 20, 6, 17 samples, respectively.
7. **Lymphoma** [110] is a multi-class dataset containing 96 samples with 4026 gene expressions as features. The gene expression profiles correspond to the three most prevalent adult lymphoid malignancies: Diffuse Large B-Cell Lymphoma (DL-BCL), Follicular Lymphoma (FL), and B-Cell Chronic Lymphocytic Leukemia (B-CLL).
8. **Prostate Tumor** dataset [111] is collected from 52 tumor samples and 50 normal samples (i.e., 102 samples in total), where each sample has 10509 gene expressions as features.
9. **Small Round Blue Cell Tumors (SRBCT)** dataset [112] is a multi-category, gene expression dataset of 83 samples, 2308 gene expressions as features, and four class labels. 8) Prostate Cancer dataset is collected from 52 tumor samples and 50 normal samples (i.e., 102 samples in total), where each sample has 10509 gene expressions as features.
10. **TOX-171** [21] is a gene expression dataset with 5748 gene expressions as features in 171 patients with various skin conditions. There are four categories of patients in the dataset: suffered radiation-therapy (RadS) patients, controlled radiation-therapy (RadC) patients, patients with skin cancer (SkCa), and patients with no cancer (NoCa).

### 4.2 Experimental Setting

To run our experiments, we created a pipeline that consisted of two components: feature selection and clustering. Each dataset was passed through the first component to select a subset of the initial features by using a feature selection algorithm. Then, we ran the dataset with selected features through a k-means clustering model as the downstream task [113] to evaluate the effectiveness of the feature selection algorithm in separating samples. We needed to specify some parameters in order to run the feature selection algorithms and also the k-means model as the downstream task. For all feature selection algorithms and all datasets, we searched the number of the selected features *k* from the set {10^*t*^ | *t* = 1, …, 10}. Furthermore, we fixed the number of maximum iterations as 30 for the feature selection models. The penalty parameter *ρ* for the methods MFFS, MPMR, SGFS and SLSDR was searched in {10^*t*^ | *t* = − 3, …, 8}. Additionally, the redundancy parameter for MPMR was set to 1. The sparsity regularization parameter for RMFFS and SLSDR was chosen from {10^*t*^ | *t* = 0, …, 8}. Finally, the other regularization parameters for SGFS and SLSDR were tuned from {10^*t*^ | *t* = − 8, …, 8}. For SGFS and SLSDR, the k-nearest neighborhood method was utilized to construct the weighted matrix in which the size of neighbors was set from {3, 5, 10}. Moreover, the bandwidth parameter *σ* in the Gaussian kernel was selected within the range {10^*t*^ | *t* = 0, …, 6}.

Since k-means clustering is sensitive to the initial random values of the centroids, we repeated the clustering task on all gene expression datasets 20 times. Then, we calculated some statistics, as explained below, to evaluate the clustering performance. We tuned the parameters of the feature selection algorithms to obtain the best clustering metrics. We should note that we set the number of clusters in k-means clustering to be the number of class labels in the datasets.

### 4.3 Evaluation Metrics for Comparison

We selected the Clustering Accuracy (ACC) and Normalized Mutual Information (NMI) as the evaluation metrics for the clustering model [22]. These evaluation metrics are defined below:

- **ACC:** It gives us the percentage of ground truth labels that are correctly predicted by the clustering algorithm and is calculated as

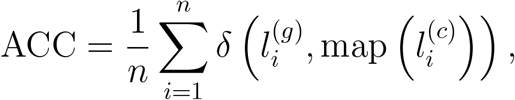

where 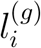 and 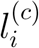 are the ground truth and clustering labels for the *i*th data point and *n* is the total number of data points. The *δ*(·,·) function is an indicator function which evaluates to one for identical inputs to the function and is equal to zero otherwise. The map(·) function maps the clustering label 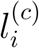 to the corresponding label of the dataset.
- **NMI:** This metric is defined for two random variables *p* and *q* as

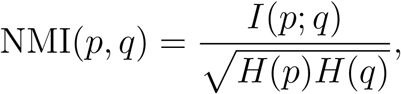

where *I*(·) and *H*(·) represent the mutual information and the entropy of the input data, respectively. We use NMI to measure the quality of clustering, where the higher values of NMI implies the better clustering performance. If we consider the predicted labels by clustering model as 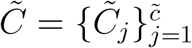 and the true labels of clusters as 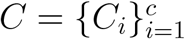, then NMI can be expressed as follows:

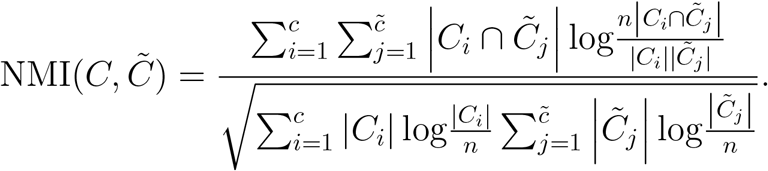

### 4.4 Results and Discussion

We analyze the performance of the feature selection algorithms in this section based on the performance of clustering models applied to the gene expression datasets with the selected subset of features.

All feature selection techniques are compared to each other in Figure 2 and Figure 3 based on ACC and NMI clustering metrics, respectively. The numerical values of the metrics are also presented in Tables 3 and 4, respectively. The Baseline in these figures and tables corresponds to the case where the k-means clustering was applied to datasets with their original features.

**Table 3:**
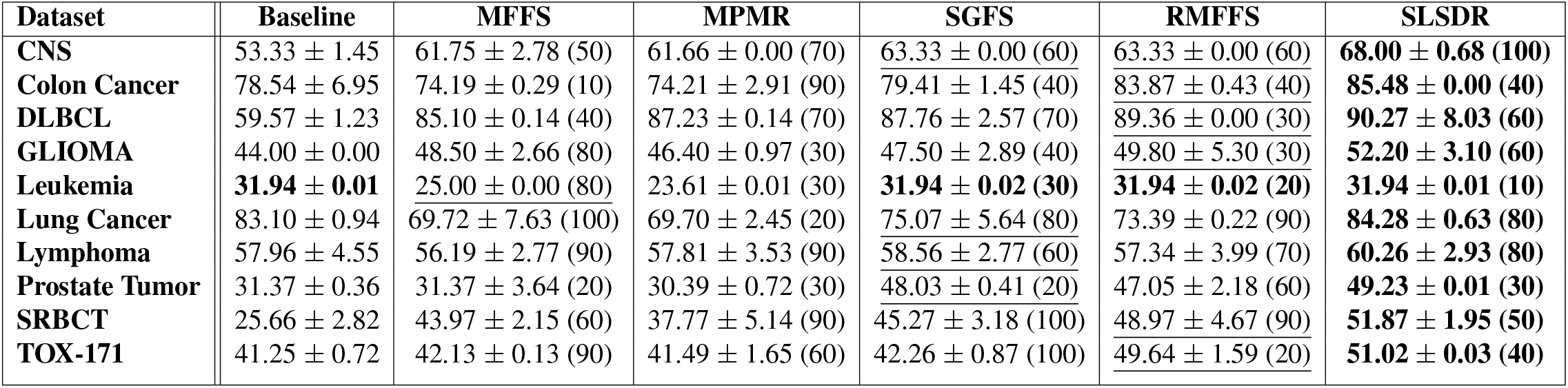
The results of clustering accuracy (ACC±STD%) corresponding to five feature selection techniques computed on seven datasets. In every row, the first and the second best outcomes are boldfaced and are underscored, respectively. The number of the selected features for the best clustering outcomes is shown in parentheses. (A bigger value of ACC indicates a better clustering performance.)

| Dataset | Baseline | MFFS | MPMR | SGFS | RMFFS | SLSDR |
| --- | --- | --- | --- | --- | --- | --- |
| CNS | 53.33 ± 1.45 | 61.75 ± 2.78 (50) | 61.66 ± 0.00 (70) | 63.33 ± 0.00 (60) | 63.33 ± 0.00 (60) | **68.00 ± 0.68 (100)** |
| Colon Cancer | 78.54 ± 6.95 | 74.19 ± 0.29 (10) | 74.21 ± 2.91 (90) | 79.41 ± 1.45 (40) | 83.87 ± 0.43 (40) | **85.48 ± 0.00 (40)** |
| DLBCL | 59.57 ± 1.23 | 85.10 ± 0.14 (40) | 87.23 ± 0.14 (70) | 87.76 ± 2.57 (70) | 89.36 ± 0.00 (30) | **90.27 ± 8.03 (60)** |
| GLIOMA | 44.00 ± 0.00 | 48.50 ± 2.66 (80) | 46.40 ± 0.97 (30) | 47.50 ± 2.89 (40) | 49.80 ± 5.30 (30) | **52.20 ± 3.10 (60)** |
| Leukemia | **31.94 ± 0.01** | 25.00 ± 0.00 (80) | 23.61 ± 0.01 (30) | **31.94 ± 0.02 (30)** | **31.94 ± 0.02 (20)** | **31.94 ± 0.01 (10)** |
| Lung Cancer | 83.10 ± 0.94 | 69.72 ± 7.63 (100) | 69.70 ± 2.45 (20) | 75.07 ± 5.64 (80) | 73.39 ± 0.22 (90) | **84.28 ± 0.63 (80)** |
| Lymphoma | 57.96 ± 4.55 | 56.19 ± 2.77 (90) | 57.81 ± 3.53 (90) | 58.56 ± 2.77 (60) | 57.34 ± 3.99 (70) | **60.26 ± 2.93 (80)** |
| Prostate Tumor | 31.37 ± 0.36 | 31.37 ± 3.64 (20) | 30.39 ± 0.72 (30) | 48.03 ± 0.41 (20) | 47.05 ± 2.18 (60) | **49.23 ± 0.01 (30)** |
| SRBCT | 25.66 ± 2.82 | 43.97 ± 2.15 (60) | 37.77 ± 5.14 (90) | 45.27 ± 3.18 (100) | 48.97 ± 4.67 (90) | **51.87 ± 1.95 (50)** |
| TOX-171 | 41.25 ± 0.72 | 42.13 ± 0.13 (90) | 41.49 ± 1.65 (60) | 42.26 ± 0.87 (100) | 49.64 ± 1.59 (20) | **51.02 ± 0.03 (40)** |

**Table 4:** The results of Normalized mutual information (NMI±STD%) corresponding to five feature selection techniques computed on seven datasets. In every row, the first and the second best outcomes are boldfaced and are underscored, respectively. The number of the selected features for the best clustering outcomes is shown in parentheses. (A bigger value of NMI indicates a better clustering performance.)

| Dataset | Baseline | MFFS | MPMR | SGFS | RMFFS | SLSDR |
| --- | --- | --- | --- | --- | --- | --- |
| CNS | $1.17 \pm 0.00$ | $2.93 \pm 0.45$ (100) | $1.60 \pm 0.80$ (30) | $8.79 \pm 0.00$ (30) | $9.35 \pm 0.08$ (50) | **<u>$9.94 \pm 3.66$</u>** (100) |
| Colon Cancer | $27.35 \pm 9.96$ | $24.30 \pm 0.14$ (100) | $21.08 \pm 0.54$ (40) | $30.03 \pm 0.00$ (10) | $33.55 \pm 0.04$ (60) | **<u>$41.58 \pm 0.03$</u>** (40) |
| DLBCL | $2.93 \pm 1.09$ | $39.45 \pm 0.00$ (40) | $48.46 \pm 0.72$ (70) | $49.51 \pm 0.72$ (70) | $55.50 \pm 0.65$ (30) | **<u>$60.70 \pm 1.24$</u>** (60) |
| GLIOMA | $17.94 \pm 0.69$ | $23.69 \pm 2.07$ (80) | $25.24 \pm 2.08$ (30) | $27.59 \pm 6.86$ (90) | $29.81 \pm 1.46$ (30) | **<u>$32.42 \pm 3.01$</u>** (100) |
| Leukemia | $21.47 \pm 0.00$ | $66.56 \pm 0.06$ (80) | $62.07 \pm 0.03$ (30) | $72.87 \pm 0.00$ (30) | $80.05 \pm 0.01$ (70) | **<u>$83.78 \pm 0.03$</u>** (30) |
| Lung Cancer | $68.12 \pm 0.56$ | $55.61 \pm 6.88$ (100) | $55.77 \pm 2.11$ (70) | $65.71 \pm 6.39$ (100) | $59.71 \pm 2.31$ (70) | **<u>$69.48 \pm 0.48$</u>** (70) |
| Lymphoma | $69.55 \pm 3.41$ | $63.47 \pm 3.03$ (60) | $67.13 \pm 3.12$ (90) | $68.93 \pm 3.12$ (60) | $65.19 \pm 2.29$ (80) | **<u>$69.50 \pm 1.90$</u>** (80) |
| Prostate Tumor | $8.56 \pm 0.21$ | $6.09 \pm 0.91$ (60) | $8.83 \pm 0.95$ (20) | $9.45 \pm 0.03$ (80) | $11.50 \pm 0.08$ (40) | **<u>$12.83 \pm 0.02$</u>** (30) |
| SRBCT | $11.55 \pm 3.47$ | $38.41 \pm 7.38$ (50) | $24.27 \pm 5.26$ (40) | $48.37 \pm 1.21$ (10) | $47.82 \pm 4.13$ (90) | **<u>$50.33 \pm 2.72$</u>** (50) |
| TOX-171 | $13.54 \pm 0.23$ | $12.82 \pm 1.52$ (80) | $16.22 \pm 1.89$ (60) | $13.96 \pm 1.02$ (100) | $24.60 \pm 0.41$ (20) | **<u>$29.90 \pm 0.69$</u>** (10) |

**Figure 2:**
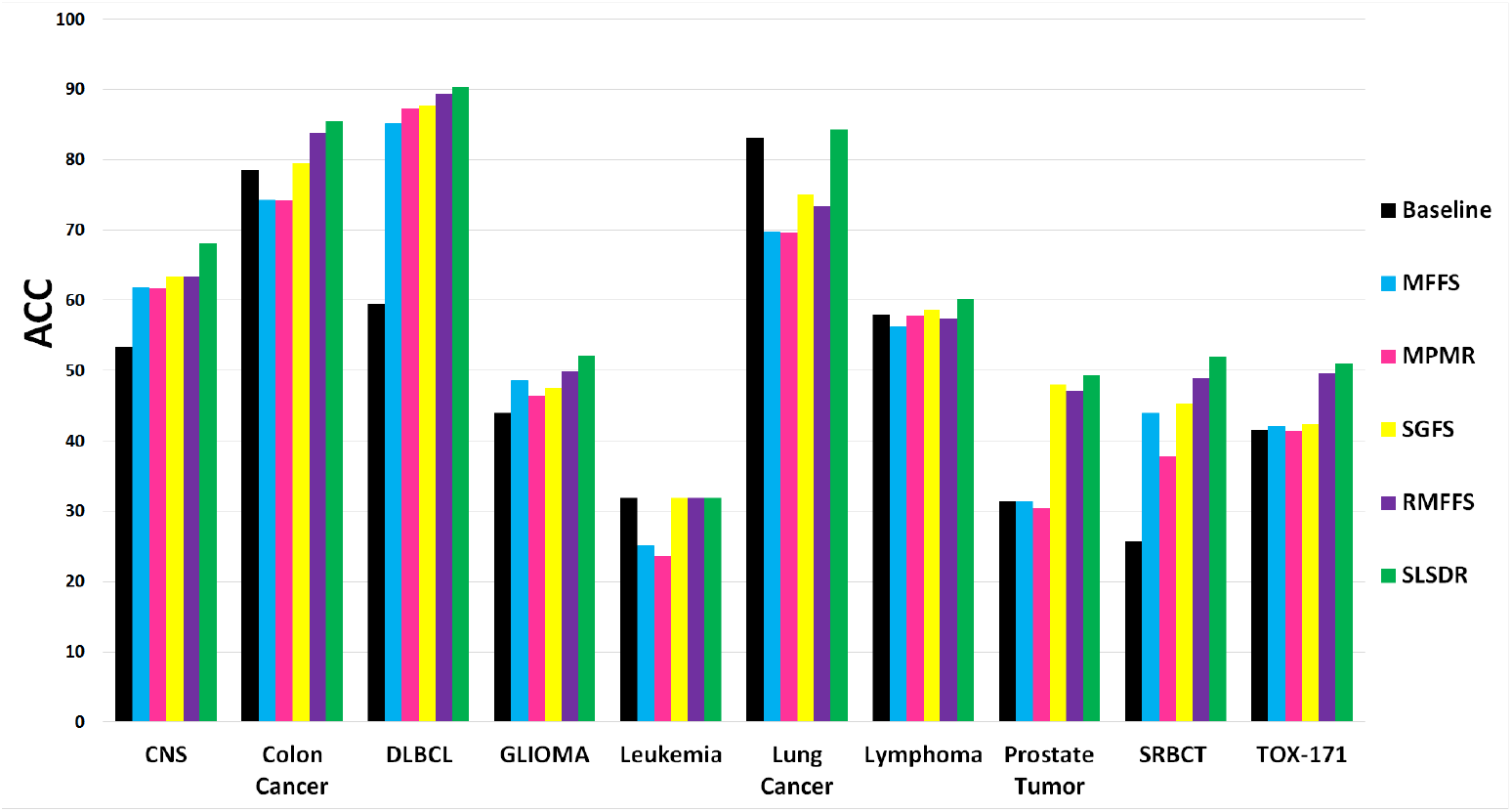
The ACC average values (the *y*-axis) versus the seven datasets (the *x*-axis). (A bigger value of ACC indicates a better clustering performance.)

**Figure 3:**
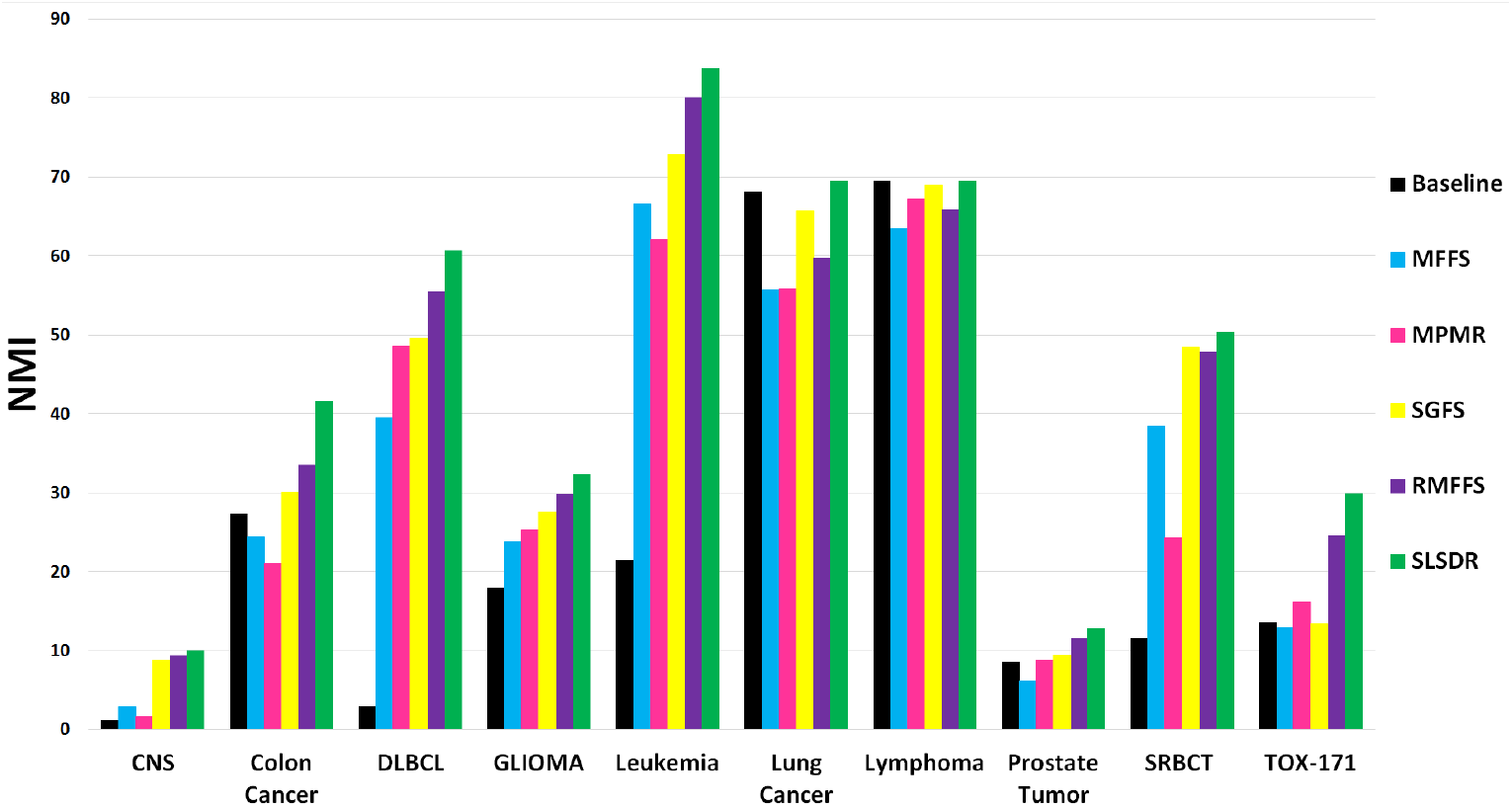
The NMI average values (the *y*-axis) versus the seven datasets (the *x*-axis). (A bigger value of NMI indicates a better clustering performance.)

Both ACC and NMI results in Figure 2 and Figure 3 clearly show the superiority of SLSDR feature selection algorithm over other algorithms in separating data points into distinct clusters for the majority of datasets. However, a closer inspection of Figure 2 and Figure 3 reveal that the influence of the selected features by SLSDR on the clustering performance differ across different datasets.

Based on the difference between SLSDR and Baseline ACCs, we can categorize the effect of feature selection by SLSDR on the clustering quality of various datasets into three levels: weak, intermediate, and strong levels. Leukemia, Lung Cancer, and Lymphoma datasets belong to the weak influence level, since the mentioned difference in ACC of clustering of these datasets is below 5%. We observe an intermediate positive effect of SLSDR on the clustering quality of Colon Cancer, GLIOMA, and TOX-171 datasets. For these datasets, the difference between ACC of SLDR and the Baseline is in the range of 5%-15%. Finally, the strong level constitutes CNS, DLBCL, Prostate Tumor, and SRBCT datasets of which the difference between ACC of SLSDR and the Baseline are above 15%. In particular, applying SLSDR to the initial feature set of DLBCL and SRBCT datasets leads to roughly 30% and 26% higher clustering ACC with respect to the Baseline feature set.

For the Leukemia dataset, neither SLSDR nor other feature selection algorithms could find features that would result in a higher clustering ACC with respect to the Baseline (i.e., clustering based on the original feature set). On the contrary, NMI value corresponding to SLSDR is almost four times larger than that of the Baseline for the Leukemia dataset.

In contrast to the Leukemia dataset, neither Lung Cancer nor Lymphoma datasets benefited from any of the feature selection schemes in terms of the clustering performance. Although SLSDR resulted in better ACC and NMI scores compared with other feature selection methods, those scores were almost identical or close to corresponding values of the Baseline.

The superior performance of SLSDR over other feature selection methods in effective clustering of some datasets can be explained by the dual-manifold aspect of SLSDR. That is, the information extracted from the geometry structures of the feature and the data manifolds at the same time enables SLSDR to obtain a rich knowledge about the local neighborhood of features. This in turn leads to a more efficient elimination of redundant features from the original dataset by SLSDR.

Considering other feature selection methods, the performance of SGFS and RMFFS is much better than that of MFFS and MPMR with respect to both clustering ACC and NMI metrics in almost all cases. For this reason, it can be deduced that the inner product regularization used in RMFFS leads to better performance in the feature selection process compared to the redundancy term used in MPMR and the orthogonality constraint used in MMFS and MPMR. Moreover, the use of the manifold regularization based on the feature space seems remarkably beneficial to raise the effectiveness level of the SGFS method.

Despite the points made about SGFS and RMFFS above, the experimental results do not support the absolute superiority of one method over another. For example, in terms of ACC, these two methods work almost identically in some cases such as CNS and Leukemia. However, in most cases, the RMFFS method outperforms SGFS which can be explained as follows. Thus, it can be inferred that the inner product regularization in RMFFS versus the feature manifold regularization in SGFS can have a better effect to eliminate redundant features in favor of the informative ones.

In Figure 4, we have presented the average clustering’s ACC and NMI scores over all gene expression datasets for different methods of feature selection methods. On average, all feature selection methods select a subset of features that results in a better clustering performance compared with the Baseline case. The clustering metrics slightly decrease when we switch from MFFS to MPMR. Then, it increases again by changing the feature selection method from MPMR to SGFS. The increasing trend in clustering performance continues as we move towards RMFFS and then SLSDR.

**Figure 4:**
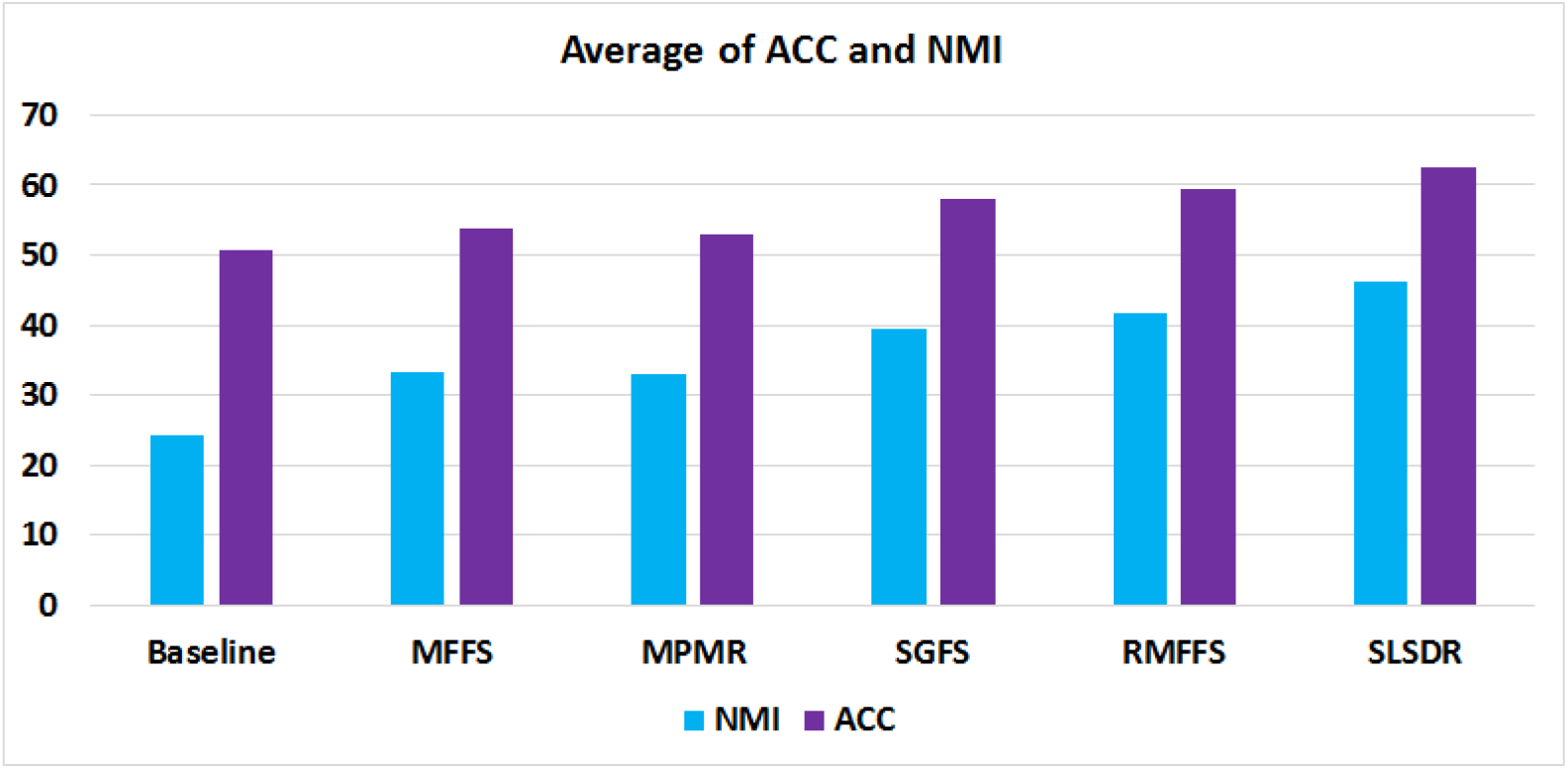
The ACC and NMI average values (the *y*-axis) versus six feature selection methods (the *x*-axis). (A bigger value of ACC or NMI indicates that the corresponding method has a better performance.)

### 4.5 Statistical Analysis

In the previous sub-section, we showed that SLSDR has superior average ACC and NMI scores compared with other feature selection techniques. In this sub-section, we try to show how statistically significant the mentioned differences are. We first consider the non-parametric Friedman test applied to the average values of ACC and NMI metrics over all datasets. This test provides a ranking of all feature selection methods based on a null hypothesis that states all of these methods lead to similar results without any significant differences. We used the Holm’s procedure as a post-hoc analysis in order to verify the differences observed among these methods.

We have demonstrated the average rankings of various feature selection methods in Figure 5 that are obtained by the Friedman test based on the average ACC and NMI scores. Methods with lower ranks possess higher performance. Therefore, SLSDR and RMFFS have, respectively, the first- and second-best performance in terms of both clustering ACC and NMI scores. The ACC ranking of MPMR is a bit higher than that of the Baseline, which indicates that a dataset whose feature set is selected by MPMR would potentially have lower clustering ACC score compared to the Baseline. The ACC results in Figure 2 for the Colon Cancer, Leukemia, Lung Cancer, and Prostate Tumor datasets agree well with the this conclusion.

**Figure 5:**
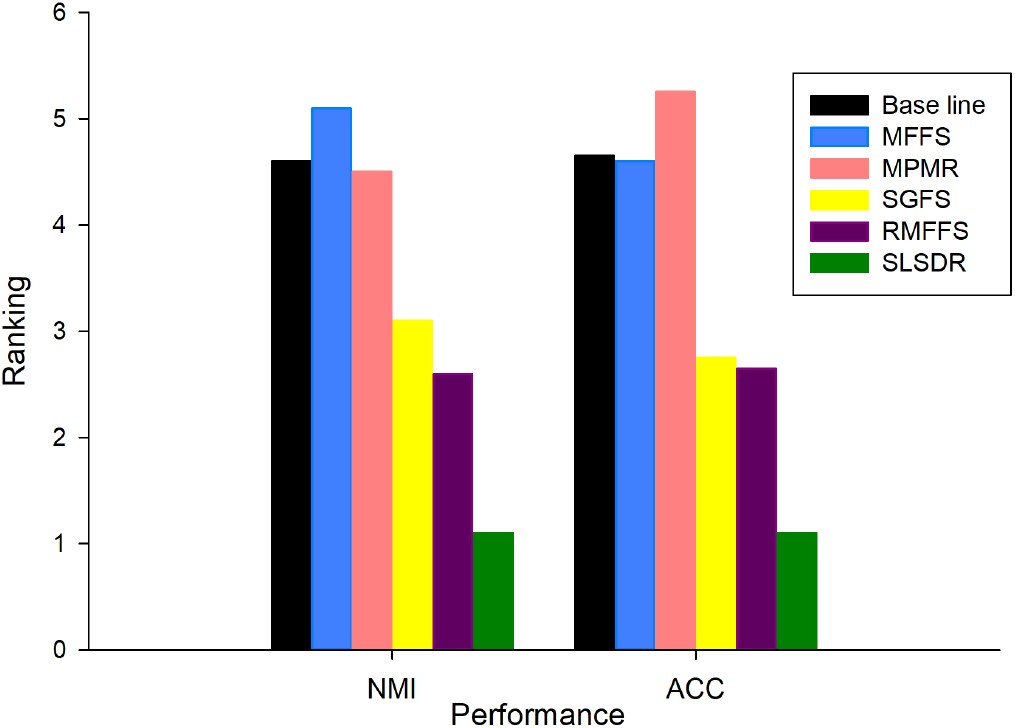
Average ranks obtained by the Friedman test for each method with respect to the different evaluation metrics on the datasets. (The lower rank of the evaluation metrics, the better the performance of the methods.)

We also performed the Holm’s procedure to do pairwise comparisons between methods to infer any statistically significant difference between them in terms of clustering metrics. The results are presented in Table 5 and Table 6 for ACC and NMI metrics respectively. We have set SLSDR as the control method and the significance level *α* to be 0.05. For ACC and NMI cases, if the Holm’s *p*-value of a pairwise comparison is less than or equal to 0.025, the Holm’s procedure rejects the null hypothesis. Table 5 clearly shows that the difference between the clustering ACC of SLSDR method on the one hand and that of the Baseline, MPMR, MFFS, and SGFS on the other hand is statistically significant. A similar conclusion can be drawn from Table 6 based on the NMI metric. Because the Holm’s *p*-values of these four methods are *<*= 0.025 for both ACC and NMI cases. However, the Holm’s procedure fails to reject the null hypothesis for RMFFS method, since its corresponding Holm’s *p*-value is greater than 0.025. In other words, it is safe to infer from the Holm’s procedure results that there is no statistically significant difference between SLSDR and RMFFS methods from either of clustering ACC or NMI perspectives.

**Table 5:** Post hoc comparisons on the ACC metric by using the significance level *α* = 0.05. Here, the control method is SLSDR, and the Holm’s procedure rejects a null hypothesis when the Holm’s *p*-value of a pairwise comparison ≤ 0.025.

| Method | Holm’s $p$-value | Reject |
| --- | --- | --- |
| MPMR | 0.01 | Yes |
| Baseline | 0.0125 | Yes |
| MFFS | 0.016667 | Yes |
| SGFS | 0.025 | Yes |
| RMFFS | 0.05 | No |

**Table 6:** Post hoc comparisons on the NMI metric by using the significance level *α* = 0.05. Here, the control method is SLSDR, and the Holm’s procedure rejects a null hypothesis when the Holm’s *p*-value of a pairwise comparison ≤ 0.05.

| Method | Holm’s $p$-value | Reject |
| --- | --- | --- |
| MFFS | 0.01 | Yes |
| Baseline | 0.0125 | Yes |
| MPMR | 0.016667 | Yes |
| SGFS | 0.025 | Yes |
| RMFFS | 0.05 | Yes |

### 4.6 Computational Complexity Analysis

The superior feature selection performance of SLSDR comes at a high computational cost compared to other methods. Table 7 compares the per-iteration computational complexity of different feature selection algorithms in this work. The SLSDR’s computational complexity, *θ*^SLSDR^, is different from that of other methods in two ways. First, *θ*^SLSDR^ is a quadratic function of number of samples (*n*), whereas the computational complexities of MFFS, MPMR, and RMFFS are independent of *n*. The SGFS’s computational complexity is also a function of *n*, yet it is a linear dependency. Thus, in the worst-case scenario, when *n* is on the same order of the number of features, *d*, the time complexity of SLSDR becomes cubic (*O*(*n*^3^)) in a large, high dimensional dataset. Second, as opposed to other methods, *θ*^SLSDR^ is also a function of the number of selected features (*k*). Figure 6 illustrates the runtime of various feature selection methods for different gene expression datasets as a function of *k*, where the value of *k* is selected from the set {10, 40, 80, 100}. It is evident from Figure 6 that SLSDR has substantially longer runtime than other methods. Further, as the number of selected features increases from 10 to 100, so does the runtime.

**Table 7:** The per-iteration computational complexity comparison among different feature selection methods. Note that *n* is the number of samples, *d* is the number of features, and *k* is the number of selected features.

| Method | Computational complexity |
| --- | --- |
| **MFFS** | $O(d^2)$ |
| **MPMR** | $O(d^2)$ |
| **SGFS** | $O(d^2) + O(nd^2)$ |
| **RMFFS** | $O(d^2)$ |
| **SLSDR** | $O((k + d)(n^2 + nd + kd)) + O(nd^2 + dn^2)$ |

**Figure 6:**
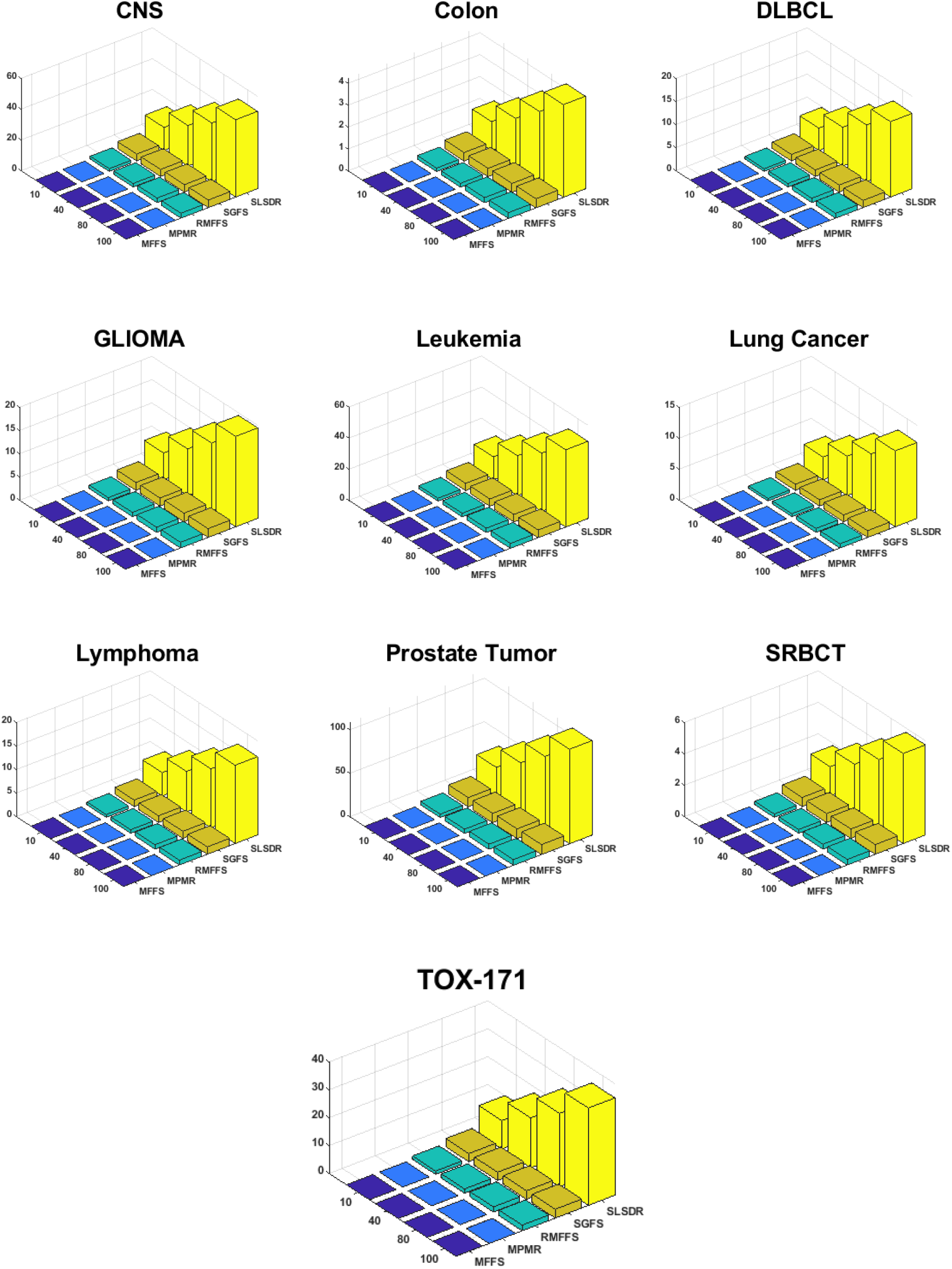
The runtime of various feature selection methods for different gene expression datasets as a function of *k*, where the value of *k* ∈ {10, 40, 80, 100}.

## 5 Application to a COVID-19 clinical dataset

In this section, we evaluate the performance of the feature selection algorithms on classifying whether patients who have COVID-19 survive or not. The COVID-19 clinical dataset was collected at Birjand University of Medical Sciences from March 2020 to August 2020 and includes clinical data from 500 patients and 66 blood clinical markers. The COVID-19 diagnosis in these patients was confirmed by a positive PCR-based clinical laboratory testing for SARS-CoV-2*.

Due to the relatively small number of samples in the COVID-19 clinical dataset, any machine learning model that is trained on the full set of features is prone to overfitting. To resolve this issue, we trained the classifier on a subset of features that are less correlated with each other and more predictive of the class labels.

Since Random Forest algorithm is generally robust against overfitting [114], we decided to use it to train a classification model on the COVID-19 dataset. To control for overfitting and generalizability to unseen data, we adapted a special training scheme that involved two nested cross-validation (CV) procedures. The outer CV uses 10 folds where the whole data is randomly partitioned to 10 subsets (folds). One subset is held out for testing and the remaining 9 subsets are joined and passed along to the inner CV. This process is repeated 10 times, each time a distinct subset is selected for testing until all 10 folds are exhausted. We used a 5-fold inner CV for training the feature selection algorithms and hyperparameter tuning.

The outcome of the inner CV is the feature selection model with optimum hyperparameters that gives us the best subset of features that can be used to effectively separate samples with different class labels. In each outer CV iteration, the Random Forest classifier is trained based on the subset of features selected by the inner CV. After the last iteration of the outer CV, the overall classification metric is obtained by averaging the performance metrics of each of the 10 Random Forest classifiers. The feature selection algorithms and their hyperparameters are the same as those mentioned in in Subsection 4.2 To investigate the effect of number of features on the classifier’s performance, we trained different classifiers for different number of features which assumed values in the range of 2, 4, 6, 8, and 10.

We employed five different metrics to evaluate the classification performance of the Random Forest model including Classification Accuracy (ACC), True Positive Rate (TPR), True Negative Rate (TNR), Positive Predictive Value (PPV), and Negative Predictive Value (NPV) [115]. The description of these classification metrics is given in the Supplementary Material. Here, it should be mentioned that when a binary classifier predicts the class label of an observation to be “Positive” or “Negative”, the predicted label can be “True” or “False” with respect to the actual (ground-truth) label of the observation. In our COVID-19 dataset, positive and negative labels correspond to death and survival conditions, respectively.

Figure 7 shows the classification metrics of the Random Forest classifier for different feature selection algorithms and different number of selected subset of features *k* = {2, 4, 6, 8, 10}. The numerical values of the metrics are also presented in Table S.1 of the Supplementary Material.

**Figure 7:**
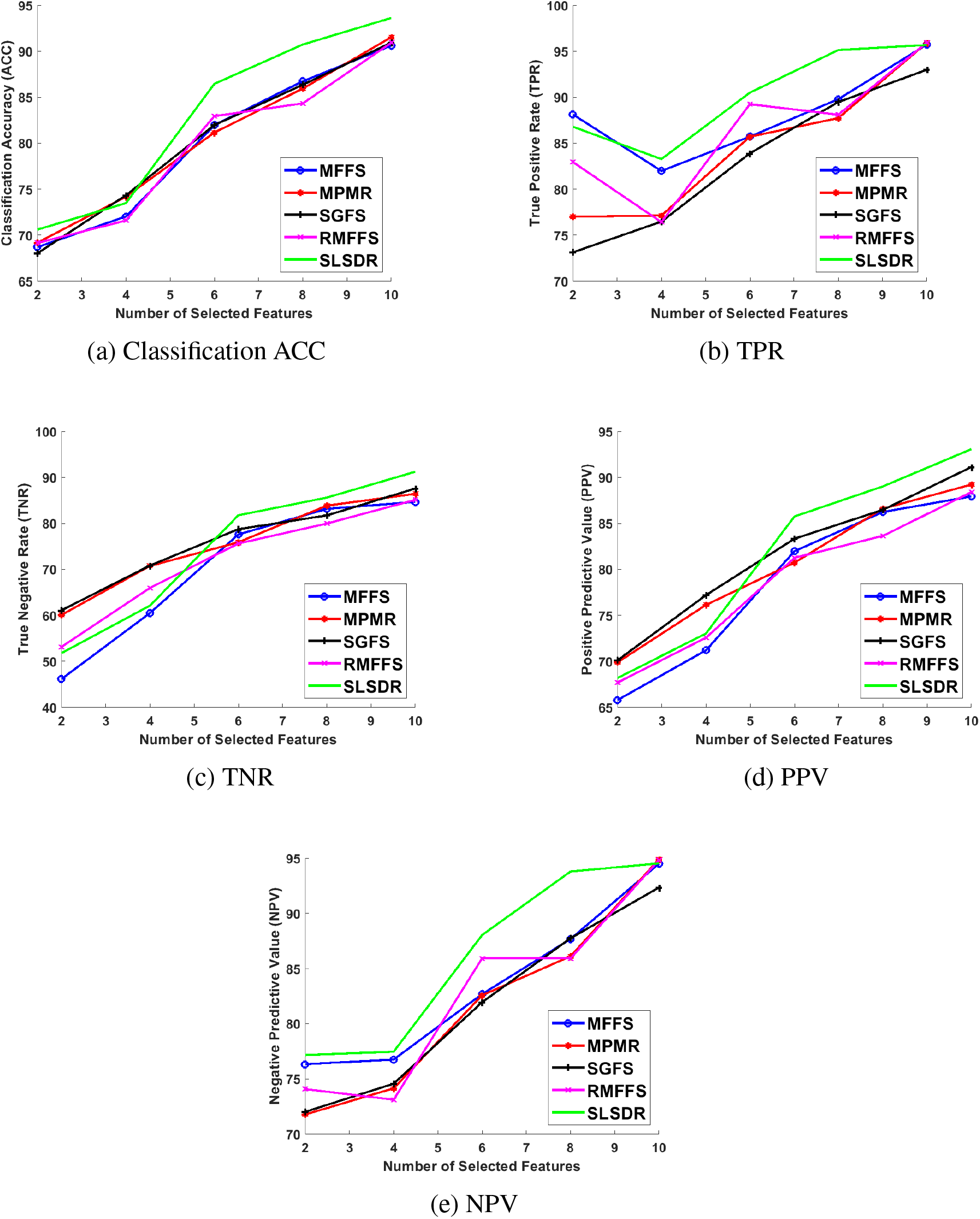
Performance metrics of the Random Forest classifier: (a) Classification ACC, (b) TPR, (c) TNR, (d) PPV, and (e) NPV.

A common theme can be readily identified in all plots of Figure 7. As *k* increases beyond 4 and more features are involved, SLSDR outperforms other feature selection algorithms in selecting a subset of features that lead to better classification metrics. We ascribe this behavior of SLSDR to its dual-manifold nature, which other methods lack. In other words, SLDR uses the geometry structures of the feature and the data manifolds at the same time such that this rich geometry information of the dataset makes SLSDR superior in comparison to the other methods. Indeed, the underlying graph network of SLSDR, which connects features, and its associated graph Laplacian matrix facilitate the search for the least redundant features that can effectively represent the original dataset.

Considering four other methods (i.e., MFFS, MPMR, SGFS and RMFFS), it is hard to assert the absolute superiority of one method over another. For example, in terms of the classification ACC in Figure 7a, these four methods result in almost similar classification performance. In terms of TPR (see Figure 7b), the MFFS method works better than the others for *k* = 2 and 8, whereas the RMFFS method outperforms the other methods for *k* = 6. In terms of TNR (see Figure 7c), except for *k* = 8, the SGFS method performs relatively better than other methods.

It is worth analyzing how feature selection algorithms play out with False Positive (FP) results in predicting COVID-19 survival. When the Random Forest classifier is trained on only two features, as Figure 7c shows, the maximum TNR of 61.09% is achieved when the features are selected by SGFS method. However, when the number of selected features increases, SLSDR surpasses SGFS in boosting the TNR so that it attains 91.22% at *k* = 10 which is around 4% higher than the corresponding value of SGFS. Similarly, Figure 7d shows that SGFS method leads to the highest average value of 70.11% for PPV at *k* = 2, whereas SLSDR outperforms SGFS for *k* ≥ 6 and results in a classifier with the maximum average PPV of 93.07% at *k* = 10. Considering TNR and PPV, it is clear that FPs play an important role in the variation of these two metrics. We can infer from these results that the classifier trained on two features has a relatively large number of FPs. In particular, even the best PPV value of 70.11% at *k* = 2 in our analysis implies that around 30 out of 100 COVID-19 patients are falsely predicted that they would not survive. However, the number of FPs decreases as k increases so that only 7 out of 10 COVID-19 patients would be falsely predicted to not survive. This is the case when 10 features are selected by SLSDR.

It is even more important to investigate how the classifier deals with False Negative (FN) results, because it would determine the response time and the strategy to save the lives of those who would likely die due to COVID-19. In this case, the TPR (sensitivity) results in Figure 7b and NPV values in Figure 7e can help us. On the one hand, when the Random Forest classifier is trained on 2 and 8 features that are selected by SLSDR, the average TPR values are 86.79% and 95.13%, respectively. The latter is much higher compared with the values corresponding to other feature selection methods. Even 2 out of 20 features from the COVID-19 dataset, which are selected by SLSDR, enable the classifier to achieve an average NPV of 77.16%. That is, out of 100 patients that are predicted to survive from COVID-19, 78 patients will actually survive. When 8 features are selected by SLSDR, the average NPV of the classifier reaches to 93.79%. It implies that the classifier tends to predict fewer false negatives when it is trained with 8 features selected by SLSDR. At *k* = 10, the TPR and NPV average values corresponding to all feature selection methods are almost identical, except for SGFS whose TPR and NPV avearge values are roughly 2% lower than those of other methods.

These selected features and results are important for strategic and clinical decision making in centers of COVID-19 care. While ICU beds and other critical resources are limited, focusing on narrow clinical findings (2 or 8) will optimize the medical care management since patients with risk of death can be on a priority of getting critical care. The model we introduced here, especially when it is trained on 8 features selected by SLSDR, can equip the caregivers to reliably rule out the need to assign resources to COVID-19 patients who would likely survive.

While a positive PCR test of COVID-19 confirms the infection in a patient and some clinical manifestations and characteristics like age and radiological imaging can guide a clinician on decision-making but the prediction of clinical course of the disease is a complex challenge. Data from COVID-19 patients have been used to find such clinical predictors [116, 117, 118, 119].

Figure 8 illustrates the frequency of two features that are selected by five feature selection algorithms at each iteration of the outer 10-fold CV during the training of the Random Forest classifier. Interestingly, RMFFS (see Figure 8c) and SGFS (see Figure 8d) methods have the least and the most variations across different pair of selected features. Furthermore, except for SGFS, other methods have selected (O_2_ Saturation, CRP) pair of features more often compared with other pairs. In the extreme case, RMFFS has selected the (O_2_ Saturation, CRP) pair of features in all 10 iterations of the 10-fold CV. Hypoxia (Low O_2_ Saturation on ABG) and higher abnormal levels of CRP have been reported to be associated with poor prognosis of COVID-19 disease and are shown to be correlated with higher mortality rates [120, 121, 122, 123, 124, 125]. In addition, the frequency of each individual feature, that is found in Figure 8, is added up across all five feature selection methods, and the result is demonstrated in Figure 9. Apart from CRP (frequency = 39) and O_2_ Saturation (frequency = 34), Platelet Count, Creatine, and Lymphocyte Count have also been selected, however, at much lower frequencies. These last three features have also been reported as predictive markers of poor prognosis and mortality in COVID-19 patients [126, 127, 128, 129, 130, 131].

**Figure 8:**
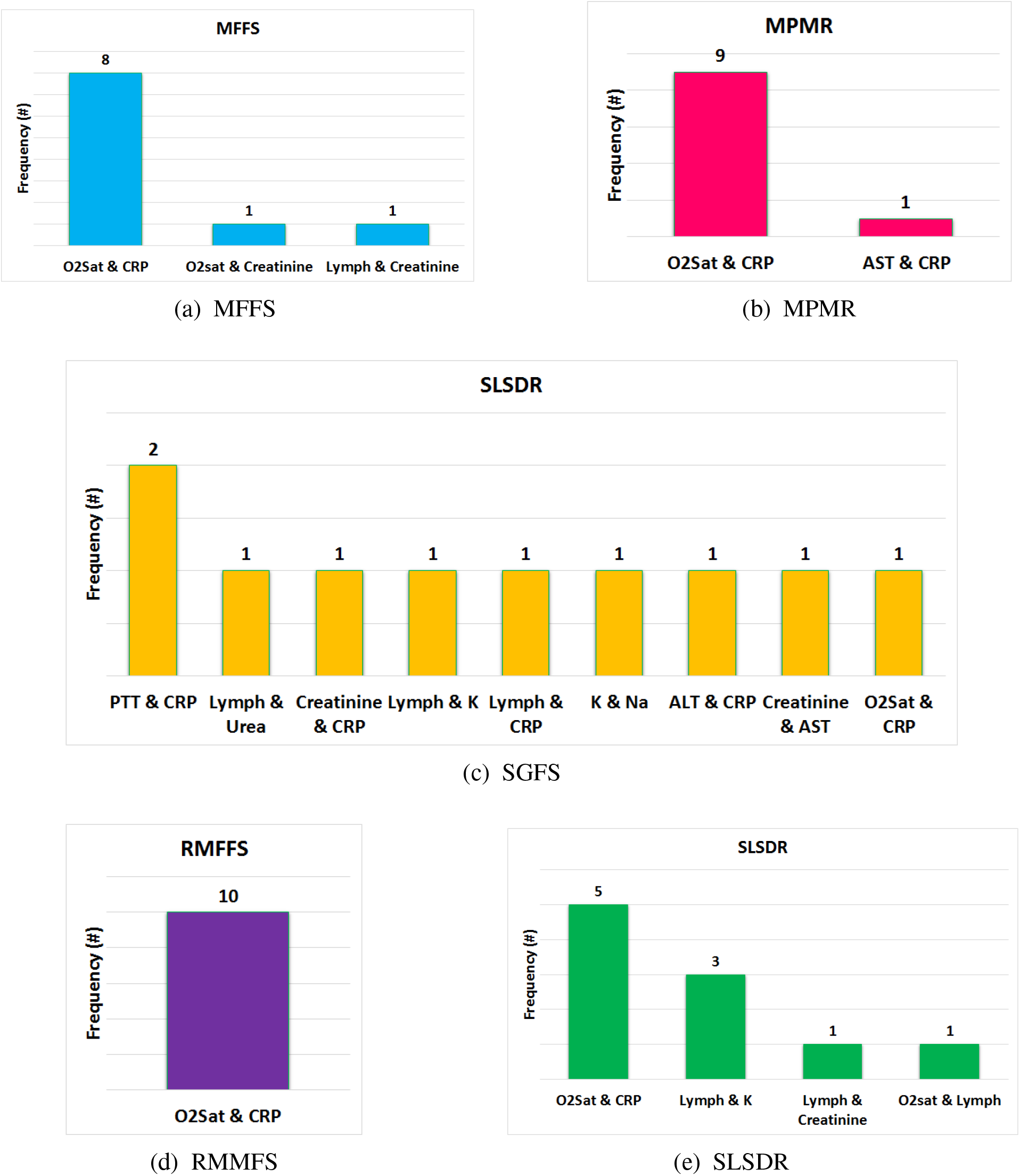
Frequency of pair of features (biomarkers) that are selected by various feature selection methods at each iteration of the 10-fold CV of the Random Forest classifier. The feature selections methods are (a) MFFS, (b) MPMR, (c) RMFSS, (d) SGFS, and (e) SLSDR.(ALT: Alanine Aminotransferase, AST: Aspartate Aminotransferase, CRP:C-Reactive Protein, K: Potassium, Lymph: Lymphocyte Count, Na: Sodium,O_2_ Sat:O_2_ Saturation on ABG, PLT: Platelet Count, PMH: Past Medical History (Cancer, Diabetes, Ischemic Heart Disease, Renal Failure, Immunodeficiency), PTT: Partial Thromboplastin Time, WBC: White Blood Cells count)

**Figure 9:**
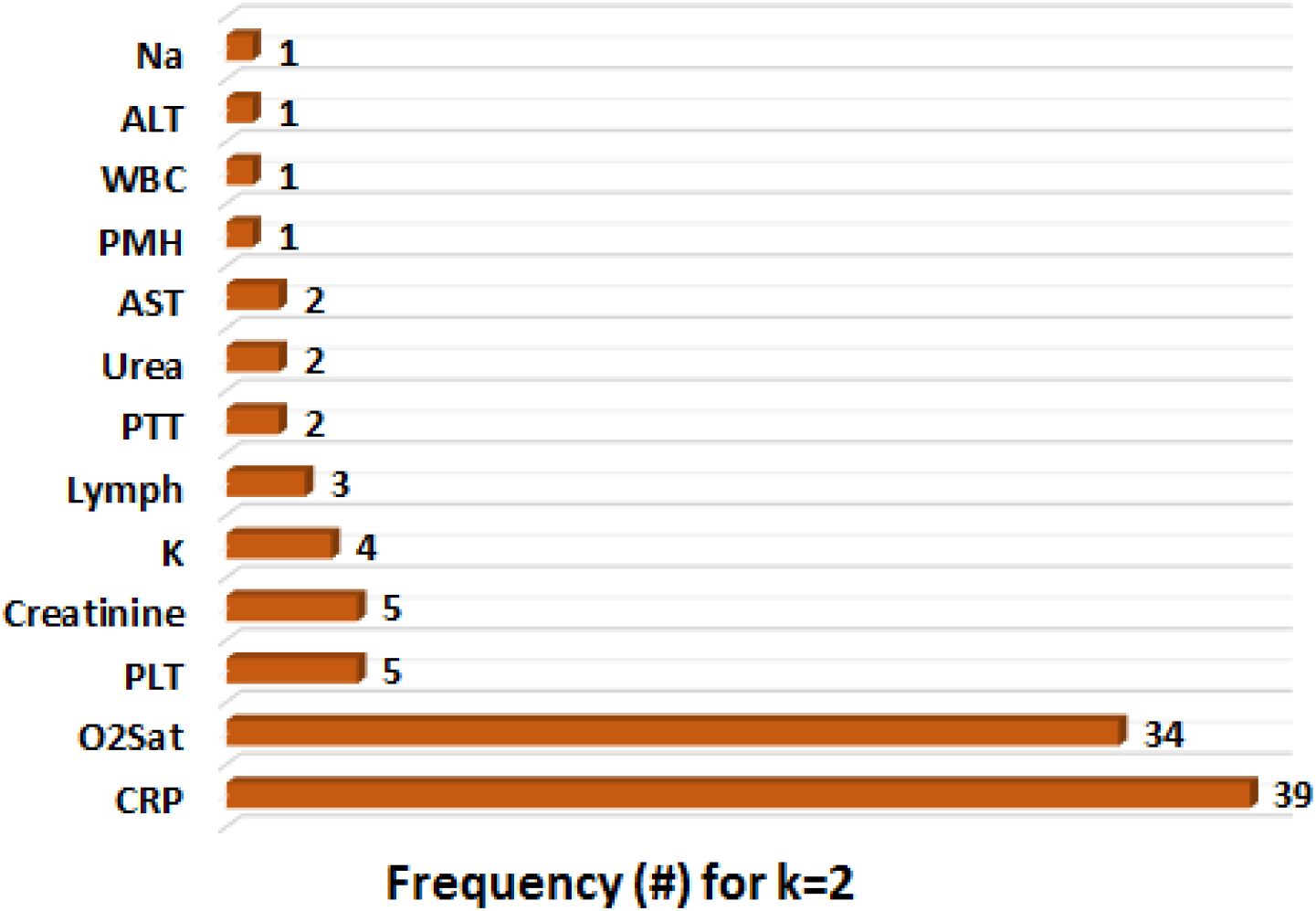
Aggregate frequency of features (biomarkers) that are selected by all feature selection methods together at all iterations of the 10-fold CV of the Random Forest classifier, where at each iteration only two features (k = 2) are selected. (ALT: Alanine Aminotransferase, AST: Aspartate Aminotransferase, CRP:C-Reactive Protein, K: Potassium, Lymph: Lymphocyte Count, Na: Sodium,O_2_ Sat:O_2_ Saturation on ABG, PLT: Platelet Count, PMH: Past Medical History (Cancer, Diabetes, Ischemic Heart Disease, Renal Failure, Immunodeficiency), PTT: Partial Thromboplastin Time, WBC: White Blood Cells count)

## 6 Conclusion

Complex diseases like COVID-19, when they appear as a pandemic, have drastic effects on the health care systems. To overcome the complications of COVID-19 on individual patients and healthcare systems, it is vital to develop advanced quantitative digital health platforms to assign the optimized clinical-decision making to each patient. To predict the prognosis of COVID-19 in a personalized approach, we need to explore the high-dimensional clinical and biomarker space of this disease to select a set of clinical and biomarker signatures. The size and content of these signatures need to be efficient in both cost and time. Although mechanistic and phenomenological predictive models in systems biomedicine [132, 133, 134, 135] are used, due to the complexity of COVID-19 and being a multi-organ disease, we need to use machine learning methodologies to reduce the high-dimensional space of clinical and biomarker spaces [13].

Using systems medicine approaches to find differentially expressed biomarkers helps explore different biomarker signatures [136]. However, essential features can be missing when applied to a clinical and biomarker space of a disease such as COVID-19. Our methodology in this paper indicates how we can discover clinical prognostic indicators for COVID-19 by reducing the high dimensionality of clinical feature space. Future clinical cohorts and systematic studies on COVID-19 can use our findings to prove the efficacy of quantitative machine learning-based clinical decision-making because is necessary to be ready for the emergence or re-emergence of other infectious diseases like COVID-19.

## Data Availability

Participants of this study did not agree for their data to be shared publicly, so supporting data is not available.

## Author Contributions

F.S.M., I.T., F.S.M., D.H. and A.M. conceived the project. M.M., M.M., F.A., E.F., S.V. and I.T. analyzed the clinical data. F.S.M., I.T., F.S.M., M.F., and A.M. designed the computational methodology. F.S.M., I.T., F.S.M., A.M., M.R.R., K.B., M,R., S.K., M.N., D.H., and M.J. did the computational analysis. I.T., M.M., M.M., E.F., F.S., and M.D., wrote the clinical section. All authors read and commented on the manuscript.

## Acknowledgments

I.T. contributed to this paper at the Cellular Energetics Program of the Kavli Institute for Theoretical Physics, supported in part by the National Science Foundation Grant NSF PHY-1748958, NIH Grant R25GM067110, and the Gordon and Betty Moore Foundation Grant 2919.02. This work was partly supported by the Intramural Research Program of the National Institute of Neurological Disorder and Stroke/ National Institutes of Health (Grant NS003031) (to F.S.).

## Abbreviations

(ALT): Alanine Aminotransferase
(ABG): Arterial Blood Gas
(AI): Artificial Intelligence
(AST): Aspartate Aminotransferase
(CNS): Central Nervous System
(ACC): Classification/Clustering Accuracy
(CBC): Complete Blood Count
(CNMF): Convex NMF
(COVID-19): Coronavirus Disease 2019
(CV): Cross-Validation
(CRP): C-Reactive Protein
(DLBCL): Diffuse large B-cell lymphoma
(FN): False Negative
(False Positive): False Positive
(FS): Feature Selection
(HSIC): Hilbert Schmidt Independence Criterion
(INR): International Normalized Ratio
(Isomap): Isometric Feature Mapping
(LDA): Linear Discriminant Analysis
(LPP): Locality Preserving Projection
(LLE): Locally Linear Embedding
(ML): Machine Learning
(MF): Matrix Factorization
(MFFS): Matrix Factorization Feature Selection
(MPMR): Maximum Projection and Minimum Redundancy
(MCFS): Multi-Cluster Feature Selection
(NPV): Negative Predictive Value
(NPE): Neighborhood Preserving Embedding
(NMF): Non-Negative Matrix Factorization
(NMI): Normalized Mutual Information
(ONMF): Orthogonal NMF
(PMH): Past Medical History
(PTT): Partial Thromboplastin Time
(PCR): Polymerase Chain Reaction
(PPV): Positive Predictive Value
(PCA): Principal Components Analysis
(PMF): Probabilistic Matrix Factorization
(RMFFS): Regularized Matrix Factorization Feature Selection
(SNMF): Semi NMF
(SVD): Singular Value Decomposition
(SRBCT): Small Round Blue Cell Tumors
SLSDR): Sparse and Low-redundant Subspace learning-based Dual-graph Regularized Robust
(SGFS): Subspace Learning-Based Graph Regularized Feature Selection
(TN): True Negative
(TNR): True Negative Rate
(TP): True Positive
(TPR): True Positive Rate
(WBC): White Blood Cells

## Footnotes

* Anonymised clinical data of all patients with COVID-19 who had been admitted in clinical centers of Birjand University of Medical Sciences during the mentioned time were used according to the Institutional Review Board (IRB) permission.

## References

[1] Johns Hopkins Coronavirus Resource Center. https://coronavirus.jhu.edu/map.html.

[2] Koichi Yuki, Miho Fujiogi, and Sophia Koutsogiannaki. COVID-19 pathophysiology: A review. Clinical Immunology, page 108427, 2020.

[3] Omid Tavassoly, Farinaz Safavi, and Iman Tavassoly. Heparin-binding peptides as novel therapies to stop SARS-CoV-2 cellular entry and infection. Molecular Pharmacology, 98(5):612–619, 2020.

[4] Omid Tavassoly, Farinaz Safavi, and Iman Tavassoly. Seeding brain protein aggregation by SARS-CoV-2 as a possible longterm complication of COVID-19 infection. ACS Chemical Neuroscience, 11(22):3704–3706, 2020.

[5] Xiaochen Li, Shuyun Xu, Muqing Yu, Ke Wang, Yu Tao, Ying Zhou, Jing Shi, Min Zhou, Bo Wu, Zhenyu Yang, et al. Risk factors for severity and mortality in adult COVID-19 inpatients in Wuhan. Journal of Allergy and Clinical Immunology, 146(1):110–118, 2020.

[6] Jonathan W Cunningham, Muthiah Vaduganathan, Brian L Claggett, Karola S Jering, Ankeet S Bhatt, Ning Rosenthal, and Scott D Solomon. Clinical outcomes in young US adults hospitalized with COVID-19. JAMA Internal Medicine, 181(3):379–381, 2021.

[7] Joseph E Ebinger, Natalie Achamallah, Hongwei Ji, Brian L Claggett, Nancy Sun, Patrick Botting, Trevor-Trung Nguyen, Eric Luong, Elizabeth H Kim, Eunice Park, et al. Preexisting traits associated with COVID-19 illness severity. PLOS One, 15(7):e0236240, 2020.

[8] Jun Mi, Weimin Zhong, Chaoqun Huang, Wanwan Zhang, Li Tan, and Lili Ding. Gender, age and comorbidities as the main prognostic factors in patients with COVID-19 pneumonia. American Journal of Translational Research, 12(10):6537, 2020.

[9] Samira Shirazi, Sanaz Mami, Negar Mohtadi, Abas Ghaysouri, Hamed Tavan, Ali Nazari, Taleb Kokhazadeh, and Reza Mollazadeh. Sudden cardiac death in COVID-19 patients, a report of three cases. Future Cardiology, 17(1):113–118, 2020.

[10] Yanjiao Lu, Zhenli Huang, Meijia Wang, Kun Tang, Shanshan Wang, Pengfei Gao, Jungang Xie, Tao Wang, and Jianping Zhao. Clinical characteristics and predictors of mortality in young adults with severe COVID-19: a retrospective observational study. Annals of Clinical Microbiology and Antimicrobials, 20(1):1–9, 2021.

[11] Brit Long, William J Brady, Alex Koyfman, and Michael Gottlieb. Cardiovascular complications in COVID-19. The American Journal of Emergency Medicine, 2020.

[12] Iman Tavassoly, Joseph Goldfarb, and Ravi Iyengar. Systems biology primer: the basic methods and approaches. Essays in Biochemistry, 62(4):487–500, 2018.

[13] Adel Mehrpooya, Farid Saberi-Movahed, Najmeh Azizizadeh, Mohammad Rezaei-Ravari, Mahdi Eftekhari, and Iman Tavassoly. High dimensionality reduction by matrix factorization for systems pharmacology. bioRxiv, 2021.

[14] Shiping Wang, Witold Pedrycz, Qingxin Zhu, and William Zhu. Subspace learning for unsupervised feature selection via matrix factorization. Pattern Recognition, 48(1):10–19, 2015.

[15] Miao Qi, Ting Wang, Fucong Liu, Baoxue Zhang, Jianzhong Wang, and Yugen Yi. Unsupervised feature selection by regularized matrix factorization. Neurocomputing, 273:593–610, 2018.

[16] Shiping Wang, Witold Pedrycz, Qingxin Zhu, and William Zhu. Unsupervised feature selection via maximum projection and minimum redundancy. Knowledge-Based Systems, 75:19–29, 2015.

[17] Ronghua Shang, Wenbing Wang, Rustam Stolkin, and Licheng Jiao. Subspace learning-based graph regularized feature selection. Knowledge-Based Systems, 112:152–165, 2016.

[18] Ronghua Shang, Kaiming Xu, Fanhua Shang, and Licheng Jiao. Sparse and low-redundant subspace learning-based dual-graph regularized robust feature selection. Knowledge-Based Systems, 187:104830, 2020.

[19] Parham Moradi and Mehrdad Rostami. A graph theoretic approach for unsupervised feature selection. Engineering Applications of Artificial Intelligence, 44:33–45, 2015.

[20] Hongmei Chen, Tianrui Li, Xin Fan, and Chuan Luo. Feature selection for imbalanced data based on neighborhood rough sets. Information Sciences, 483:1–20, 2019.

[21] Verónica Bolón-Canedo, Noelia Sánchez-Marono, Amparo Alonso-Betanzos, José Manuel Benítez, and Francisco Herrera. A review of microarray datasets and applied feature selection methods. Information Sciences, 282:111–135, 2014.

[22] Saúl Solorio-Fernández, J Ariel Carrasco-Ochoa, and José Fco Martínez-Trinidad. A review of unsupervised feature selection methods. Artificial Intelligence Review, 53(2):907–948, 2020.

[23] Girish Chandrashekar and Ferat Sahin. A survey on feature selection methods. Computers & Electrical Engineering, 40(1):16–28, 2014.

[24] Emrah Hancer, Bing Xue, and Mengjie Zhang. A survey on feature selection approaches for clustering. Artificial Intelligence Review, 53(6):4519–4545, 2020.

[25] Peter E Hart, David G Stork, and Richard O Duda. Pattern classification. John Willey & Sons, 2001.

[26] Hanchuan Peng, Fuhui Long, and Chris Ding. Feature selection based on mutual information criteria of max-dependency, max-relevance, and min-redundancy. IEEE Transactions on Pattern Analysis and Machine Intelligence, 27(8):1226–1238, 2005.

[27] Daniel D Lee and H Sebastian Seung. Learning the parts of objects by non-negative matrix factorization. Nature, 401(6755):788–791, 1999.

[28] Mehrdad Rostami, Kamal Berahmand, Elahe Nasiri, and Saman Forouzande. Review of swarm intelligence-based feature selection methods. Engineering Applications of Artificial Intelligence, 100:104210, 2021.

[29] Huan Liu and Lei Yu. Toward integrating feature selection algorithms for classification and clustering. IEEE Transactions on Knowledge and Data Engineering, 17(4):491–502, 2005.

[30] Mahdieh Labani, Parham Moradi, Fardin Ahmadizar, and Mahdi Jalili. A novel multivariate filter method for feature selection in text classification problems. Engineering Applications of Artificial Intelligence, 70:25–37, 2018.

[31] Seçkin Karasu, Aytaç Altan, Stelios Bekiros, and Wasim Ahmad. A new forecasting model with wrapper-based feature selection approach using multi-objective optimization technique for chaotic crude oil time series. Energy, 212:118750, 2020.

[32] Golnaz Sahebi, Parisa Movahedi, Masoumeh Ebrahimi, Tapio Pahikkala, Juha Plosila, and Hannu Tenhunen. GeFeS: A generalized wrapper feature selection approach for optimizing classification performance. Computers in Biology and Medicine, 125:103974, 2020.

[33] Mehrdad Rostami, Saman Forouzandeh, Kamal Berahmand, and Mina Soltani. Integration of multi-objective PSO based feature selection and node centrality for medical datasets. Genomics, 112(6):4370–4384, 2020.

[34] Di Wang, Zuoquan Zhang, Rongquan Bai, and Yanan Mao. A hybrid system with filter approach and multiple population genetic algorithm for feature selection in credit scoring. Journal of Computational and Applied Mathematics, 329:307–321, 2018.

[35] Fatemeh Aghaeipoor and Mohammad Masoud Javidi. A hybrid fuzzy feature selection algorithm for high-dimensional regression problems: An mRMR-based framework. Expert Systems with Applications, 162:113859, 2020.

[36] Gilbert Strang. Linear Algebra and Learning from Data. Cambridge Press, 2019.

[37] Charu C Aggarwal. Linear Algebra and Optimization for Machine Learning. Springer, 2020.

[38] Zafran Khan, Naima Iltaf, Hammad Afzal, and Haider Abbas. Enriching non-negative matrix factorization with contextual embeddings for recommender systems. Neurocomputing, 380:246–258, 2020.

[39] Shaily Malik and Poonam Bansal. Matrix factorization-based improved classification of gene expression data. Recent Advances in Computer Science and Communications, 13(5):858–863, 2020.

[40] Gene H Golub and Christian Reinsch. Singular value decomposition and least squares solutions. In Linear Algebra, pages 134–151. Springer, 1971.

[41] I.T. Jolliffe. Principal Component Analysis. Springer, 1986.

[42] Andriy Mnih and Russ R Salakhutdinov. Probabilistic matrix factorization. Advances in Neural Information Processing Systems, 20:1257–1264, 2007.

[43] Chong Peng, Zhilu Zhang, Zhao Kang, Chenglizhao Chen, and Qiang Cheng. Nonnegative matrix factorization with local similarity learning. Information Sciences, 562:325–346, 2021.

[44] Chris HQ Ding, Tao Li, and Michael I Jordan. Convex and semi-nonnegative matrix factorizations. IEEE Transactions on Pattern Analysis and Machine Intelligence, 32(1):45–55, 2008.

[45] Chris Ding, Tao Li, Wei Peng, and Haesun Park. Orthogonal nonnegative matrix t-factorizations for clustering. In Proceedings of the 12th ACM SIGKDD International Conference on Knowledge Discovery and Data Mining, pages 126–135, 2006.

[46] Karthik Devarajan. Nonnegative matrix factorization: an analytical and interpretive tool in computational biology. PLOS Computational Biology, 4(7):e1000029, 2008.

[47] Dong Wang, Jin-Xing Liu, Ying-Lian Gao, Chun-Hou Zheng, and Yong Xu. Characteristic gene selection based on robust graph regularized non-negative matrix factorization. IEEE/ACM Transactions on Computational Biology and Bioinformatics, 13(6):1059–1067, 2016.

[48] Andri Mirzal. Statistical analysis of microarray data clustering using NMF, spectral clustering, Kmeans, and GMM. IEEE/ACM Transactions on Computational Biology and Bioinformatics, 2020.

[49] Tayebeh Waezizadeh, Adel Mehrpooya, Maryam Rezaeizadeh, and Shantia Yarahmadian. Mathematical models for the effects of hypertension and stress on kidney and their uncertainty. Mathematical Biosciences, 305:77–95, 2018.

[50] Jin-Xing Liu, Yong Xu, Chun-Hou Zheng, Heng Kong, and Zhi-Hui Lai. RPCA-based tumor classification using gene expression data. IEEE/ACM Transactions on Computational Biology and Bioinformatics, 12(4):964–970, 2014.

[51] Suresh Balakrishnama and Aravind Ganapathiraju. Linear discriminant analysis-a brief tutorial. Institute for Signal and Information Processing, 18(1998):1–8, 1998.

[52] Ronghua Shang, Zhu Zhang, Licheng Jiao, Wenbing Wang, and Shuyuan Yang. Global discriminative-based nonnegative spectral clustering. Pattern Recognition, 55:172–182, 2016.

[53] Xiaofei He and Partha Niyogi. Locality preserving projections. Advances in Neural Information Processing Systems, 16(16):153–160, 2004.

[54] Xiaofei He, Deng Cai, Shuicheng Yan, and Hong-Jiang Zhang. Neighborhood preserving embedding. In Tenth IEEE International Conference on Computer Vision (ICCV’05) Volume 1, volume 2, pages 1208–1213. IEEE, 2005.

[55] Sam T Roweis and Lawrence K Saul. Nonlinear dimensionality reduction by locally linear embedding. Science, 290(5500):2323–2326, 2000.

[56] Georgios Tzimiropoulos, Stefanos Zafeiriou, and Maja Pantic. Subspace learning from image gradient orientations. IEEE Transactions on Pattern Analysis and Machine Intelligence, 34(12):2454–2466, 2012.

[57] Wei Zheng, Hui Yan, and Jian Yang. Robust unsupervised feature selection by nonnegative sparse subspace learning. Neurocomputing, 334:156–171, 2019.

[58] Nan Zhou, Yangyang Xu, Hong Cheng, Jun Fang, and Witold Pedrycz. Global and local structure preserving sparse subspace learning: An iterative approach to unsupervised feature selection. Pattern Recognition, 53:87–101, 2016.

[59] Cong Lei and Xiaofeng Zhu. Unsupervised feature selection via local structure learning and sparse learning. Multimedia Tools and Applications, 77(22):29605–29622, 2018.

[60] Farid Saberi-Movahed, Mahdi Eftekhari, and Mohammad Mohtashami. Supervised feature selection by constituting a basis for the original space of features and matrix factorization. International Journal of Machine Learning and Cybernetics, pages 1–17, 2019.

[61] Yanfang Liu, Dongyi Ye, Wenbin Li, Huihui Wang, and Yang Gao. Robust neighborhood embedding for unsupervised feature selection. Knowledge-Based Systems, 193:105462, 2020.

[62] Ronghua Shang, Kaiming Xu, and Licheng Jiao. Subspace learning for unsupervised feature selection via adaptive structure learning and rank approximation. Neurocomputing, 413:72–84, 2020.

[63] Joshua B Tenenbaum, Vin De Silva, and John C Langford. A global geometric framework for nonlinear dimensionality reduction. Science, 290(5500):2319–2323, 2000.

[64] Mikhail Belkin and Partha Niyogi. Laplacian Eigenmaps for dimensionality reduction and data representation. Neural Computation, 15(6):1373–1396, 2003.

[65] Xiaofei He, Deng Cai, and Partha Niyogi. Laplacian Score for feature selection. Advances in Neural Information Processing Systems, 18:507–514, 2005.

[66] Deng Cai, Chiyuan Zhang, and Xiaofei He. Unsupervised feature selection for multi-cluster data. In Proceedings of the 16th ACM SIGKDD International Conference on Knowledge Discovery and Data Mining, pages 333–342, 2010.

[67] Dong Wang, Jin-Xing Liu, Ying-Lian Gao, Chun-Hou Zheng, and Yong Xu. Characteristic gene selection based on robust graph regularized non-negative matrix factorization. IEEE/ACM Transactions on Computational Biology and Bioinformatics, 13(6):1059–1067, 2015.

[68] Yang Meng, Ronghua Shang, Licheng Jiao, Wenya Zhang, Yijing Yuan, and Shuyuan Yang. Feature selection based dual-graph sparse non-negative matrix factorization for local discriminative clustering. Neurocomputing, 290:87–99, 2018.

[69] Chang Tang, Xinwang Liu, Miaomiao Li, Pichao Wang, Jiajia Chen, Lizhe Wang, and Wanqing Li. Robust unsupervised feature selection via dual self-representation and manifold regularization. Knowledge-Based Systems, 145:109–120, 2018.

[70] Shaoyong Li, Chang Tang, Xinwang Liu, Yaping Liu, and Jiajia Chen. Dual graph regularized compact feature representation for unsupervised feature selection. Neurocomputing, 331:77–96, 2019.

[71] Qing Ye, Xiaolong Zhang, and Yaxin Sun. Dual global structure preservation based supervised feature selection. Neural Processing Letters, pages 1–23, 2020.

[72] Peng Luo, Jinye Peng, Ziyu Guan, and Jianping Fan. Dual regularized multi-view non-negative matrix factorization for clustering. Neurocomputing, 294:1–11, 2018.

[73] Chang Tang, Meiru Bian, Xinwang Liu, Miaomiao Li, Hua Zhou, Pichao Wang, and Hailin Yin. Unsupervised feature selection via latent representation learning and manifold regularization. Neural Networks, 117:163–178, 2019.

[74] Ying Ren, Min-Yu Tsai, Liyuan Chen, Jing Wang, Shulong Li, Yufei Liu, Xun Jia, and Chenyang Shen. A manifold learning regularization approach to enhance 3D CT image-based lung nodule classification. International Journal of Computer Assisted Radiology and Surgery, 15(2):287–295, 2020.

[75] Mohammad Rezaei-Ravari, Mahdi Eftekhari, and Farid Saberi-Movahed. Regularizing extreme learning machine by dual locally linear embedding manifold learning for training multi-label neural network classifiers. Engineering Applications of Artificial Intelligence, 97:104062, 2021.

[76] Rui Zhang, Yunxing Zhang, and Xuelong Li. Unsupervised feature selection via adaptive graph learning and constraint. IEEE Transactions on Neural Networks and Learning Systems, 2020.

[77] Jiawei Han, Micheline Kamber, and Jian Pei. Data mining concepts and techniques third edition. The Morgan Kaufmann Series in Data Management Systems, 5(4):83–124, 2011.

[78] Jacob Benesty, Jingdong Chen, and Yiteng Huang. On the importance of the Pearson correlation coefficient in noise reduction. IEEE Transactions on Audio, Speech, and Language Processing, 16(4):757–765, 2008.

[79] Jacob Benesty, Jingdong Chen, Yiteng Huang, and Israel Cohen. Pearson correlation coefficient. In Noise Reduction in Speech Processing, pages 1–4. Springer, 2009.

[80] Mark Andrew Hall. Correlation-based feature selection for machine learning. PhD thesis, University of Waikato Hamilton, 1999.

[81] Pabitra Mitra, CA Murthy, and Sankar K. Pal. Unsupervised feature selection using feature similarity. IEEE Transactions on Pattern Analysis and Machine Intelligence, 24(3):301–312, 2002.

[82] Michal Haindl, Petr Somol, Dimitrios Ververidis, and Constantine Kotropoulos. Feature selection based on mutual correlation. In Iberoamerican Congress on Pattern Recognition, pages 569–577. Springer, 2006.

[83] Darío García-García and Raúl Santos-Rodríguez. Spectral clustering and feature selection for microarray data. In 2009 International Conference on Machine Learning and Applications, pages 425–428. IEEE, 2009.

[84] Chun-Chao Yen, Liang-Chieh Chen, and Shou-De Lin. Unsupervised feature selection: minimize information redundancy of features. In 2010 International Conference on Technologies and Applications of Artificial Intelligence, pages 247–254. IEEE, 2010.

[85] Zheng Zhao, Lei Wang, Huan Liu, and Jieping Ye. On similarity preserving feature selection. IEEE Transactions on Knowledge and Data Engineering, 25(3):619–632, 2011.

[86] Sina Tabakhi and Parham Moradi. Relevance–redundancy feature selection based on ant colony optimization. Pattern Recognition, 48(9):2798–2811, 2015.

[87] Jiuqi Han, Zhengya Sun, and Hongwei Hao. Selecting feature subset with sparsity and low redundancy for unsupervised learning. Knowledge-Based Systems, 86:210–223, 2015.

[88] Zheng Zhao, Lei Wang, and Huan Liu. Efficient spectral feature selection with minimum redundancy. In Proceedings of the AAAI Conference on Artificial Intelligence, volume 24, 2010.

[89] Deng Cai, Chiyuan Zhang, and Xiaofei He. Unsupervised feature selection for multicluster data. In Proceedings of the 16th ACM SIGKDD International Conference on Knowledge Discovery and Data Mining, pages 333–342, 2010.

[90] Chenping Hou, Feiping Nie, Dongyun Yi, and Yi Wu. Feature selection via joint embedding learning and sparse regression. In Twenty-Second international joint conference on Artificial Intelligence. Citeseer, 2011.

[91] Hyunki Lim and Dae-Won Kim. Pairwise dependence-based unsupervised feature selection. Pattern Recognition, 111:107663, 2021.

[92] Mahdi Eftekhari, Farid Saberi-Movahed, and Adel Mehrpooya. Supervised feature selection via information gain, maximum projection and minimum redundancy. In SLAA10 Seminar Linear Algebra and Its Application, pages 29–35, 2020.

[93] Fadi Dornaika. Joint feature and instance selection using manifold data criteria: application to image classification. Artificial Intelligence Review, 54(3):1735–1765, 2021.

[94] Quanquan Gu and Jie Zhou. Co-clustering on manifolds. In Proceedings of the 15th ACM SIGKDD International Conference on Knowledge Discovery and Data Mining, pages 359–368, 2009.

[95] Mahla Mokhtia, Mahdi Eftekhari, and Farid Saberi-Movahed. Feature selection based on regularization of sparsity based regression models by hesitant fuzzy correlation. Applied Soft Computing, 91:106255, 2020.

[96] Zhenqiu Shu, Yunmeng Zhang, Peng Li, Congzhe You, Zhen Liu, Honghui Fan, and Xiao-jun Wu. Dual local learning regularized nonnegative matrix factorization and its semisupervised extension for clustering. Neural Computing and Applications, pages 1–19, 2020.

[97] Chengyi Zhang, Jianbo Yu, and Lyujiangnan Ye. Sparsity and manifold regu-larized convolutional autoencoders-based feature learning for fault detection of multivariate processes. Control Engineering Practice, 111:104811, 2021.

[98] Juncheng Hu, Yonghao Li, Wanfu Gao, and Ping Zhang. Robust multi-label feature selection with dual-graph regularization. Knowledge-Based Systems, 203:106126, 2020.

[99] Hua Wang, Feiping Nie, and Heng Huang. Globally and locally consistent unsupervised projection. In Proceedings of the AAAI Conference on Artificial Intelligence, volume 28, 2014.

[100] Jian Yang, David Zhang, Jing-yu Yang, and Ben Niu. Globally maximizing, locally minimizing: unsupervised discriminant projection with applications to face and palm biometrics. IEEE Transactions on Pattern Analysis and Machine Intelligence, 29(4):650–664, 2007.

[101] Feature Selection Datasets at Arizona State University. http://featureselection.asu.edu/datasets.php.

[102] Jundong Li, Kewei Cheng, Suhang Wang, Fred Morstatter, Robert P Trevino, Jiliang Tang, and Huan Liu. Feature selection: A data perspective. ACM Computing Surveys (CSUR), 50(6):94, 2018.

[103] Kent Ridge Biomedical Data Set Repository. https://leo.ugr.es/elvira/DBCRepository/.

[104] Scott L Pomeroy, Pablo Tamayo, Michelle Gaasenbeek, Lisa M Sturla, Michael Angelo, Margaret E McLaughlin, John YH Kim, Liliana C Goumnerova, Peter M Black, Ching Lau, et al. Prediction of central nervous system embryonal tumour outcome based on gene expression. Nature, 415(6870):436–442, 2002.

[105] Uri Alon, Naama Barkai, Daniel A Notterman, Kurt Gish, Suzanne Ybarra, Daniel Mack, and Arnold J Levine. Broad patterns of gene expression revealed by clustering analysis of tumor and normal colon tissues probed by oligonucleotide arrays. Proceedings of the National Academy of Sciences, 96(12):6745–6750, 1999.

[106] Ash A Alizadeh, Michael B Eisen, R Eric Davis, Chi Ma, Izidore S Lossos, Andreas Rosenwald, Jennifer C Boldrick, Hajeer Sabet, Truc Tran, Xin Yu, et al. Distinct types of diffuse large B-cell lymphoma identified by gene expression profiling. Nature, 403(6769):503–511, 2000.

[107] Catherine L Nutt, DR Mani, Rebecca A Betensky, Pablo Tamayo, J Gregory Cairncross, Christine Ladd, Ute Pohl, Christian Hartmann, Margaret E McLaughlin, Tracy T Batchelor, et al. Gene expression-based classification of malignant gliomas correlates better with survival than histological classification. Cancer Research, 63(7):1602–1607, 2003.

[108] Todd R Golub, Donna K Slonim, Pablo Tamayo, Christine Huard, Michelle Gaasenbeek, Jill P Mesirov, Hilary Coller, Mignon L Loh, James R Downing, Mark A Caligiuri, et al. Molecular classification of cancer: class discovery and class prediction by gene expression monitoring. Science, 286(5439):531–537, 1999.

[109] Arindam Bhattacharjee, William G Richards, Jane Staunton, Cheng Li, Stefano Monti, Priya Vasa, Christine Ladd, Javad Beheshti, Raphael Bueno, Michael Gillette, et al. Classification of human lung carcinomas by mRNA expression profiling reveals distinct adenocarcinoma subclasses. Proceedings of the National Academy of Sciences, 98(24):13790–13795, 2001.

[110] Ash A Alizadeh, Michael B Eisen, R Eric Davis, Chi Ma, Izidore S Lossos, Andreas Rosenwald, Jennifer C Boldrick, Hajeer Sabet, Truc Tran, Xin Yu, et al. Distinct types of diffuse large B-cell lymphoma identified by gene expression profiling. Nature, 403(6769):503–511, 2000.

[111] Dinesh Singh, Phillip G Febbo, Kenneth Ross, Donald G Jackson, Judith Manola, Christine Ladd, Pablo Tamayo, Andrew A Renshaw, Anthony V D’Amico, Jerome P Richie, et al. Gene expression correlates of clinical prostate cancer behavior. Cancer Cell, 1(2):203–209, 2002.

[112] Javed Khan, Jun S Wei, Markus Ringner, Lao H Saal, Marc Ladanyi, Frank Westermann, Frank Berthold, Manfred Schwab, Cristina R Antonescu, Carsten Peterson, et al. Classification and diagnostic prediction of cancers using gene expression profiling and artificial neural networks. Nature Medicine, 7(6):673–679, 2001.

[113] Arthur P Dempster, Nan M Laird, and Donald B Rubin. Maximum likelihood from incomplete data via the EM algorithm. Journal of the Royal Statistical Society: Series B (Methodological), 39(1):1–22, 1977.

[114] Leo Breiman. Random forests. Machine Learning, 45(1):5–32, 2001.

[115] Stephen V Stehman. Selecting and interpreting measures of thematic classification accuracy. Remote Sensing of Environment, 62(1):77–89, 1997.

[116] Jie Liu, Zilong Liu, Weipeng Jiang, Jian Wang, Mengchan Zhu, Juan Song, Xiaoyue Wang, Ying Su, Guiling Xiang, Maosong Ye, et al. Clinical predictors of COVID-19 disease progression and death: Analysis of 214 hospitalised patients from Wuhan, China. The Clinical Respiratory Journal, 15(3):293–309, 2021.

[117] Qiurong Ruan, Kun Yang, Wenxia Wang, Lingyu Jiang, and Jianxin Song. Clinical predictors of mortality due to COVID-19 based on an analysis of data of 150 patients from Wuhan, China. Intensive Care Medicine, 46(5):846–848, 2020.

[118] Jie Liu, Zilong Liu, Weipeng Jiang, Jian Wang, Mengchan Zhu, Juan Song, Xiaoyue Wang, Ying Su, Guiling Xiang, Maosong Ye, et al. Clinical predictors of COVID-19 disease progression and death: Analysis of 214 hospitalised patients from Wuhan, China. The Clinical Respiratory Journal, 15(3):293–309, 2021.

[119] Dhruv Patel, Vikram Kher, Bhushan Desai, Xiaomeng Lei, Steven Cen, Neha Nanda, Ali Gholamrezanezhad, Vinay Duddalwar, Bino Varghese, and Assad A Oberai. Machine learning based predictors for COVID-19 disease severity. Scientific Reports, 11(1):1–7, 2021.

[120] Bikash R Sahu, Raj Kishor Kampa, Archana Padhi, and Aditya K Panda. C-reactive protein: a promising biomarker for poor prognosis in COVID-19 infection. Clinica Chimica Acta, 509:91–94, 2020.

[121] Dominic Stringer, Philip Braude, Phyo K Myint, Louis Evans, Jemima T Collins, Alessia Verduri, Terry J Quinn, Arturo Vilches-Moraga, Michael J Stechman, Lyndsay Pearce, et al. The role of C-reactive protein as a prognostic marker in COVID-19. International Journal of Epidemiology, 50(2):420–429, 2021.

[122] Philippe Brouqui, Sophie Amrane, Matthieu Million, Sébastien Cortaredona, Philippe Parola, Jean-Christophe Lagier, and Didier Raoult. Asymptomatic hypoxia in COVID-19 is associated with poor outcome. International Journal of Infectious Diseases, 102:233–238, 2021.

[123] Amir Sadeghi, Pegah Eslami, Arash Dooghaie Moghadam, Ali Pirsalehi, Sajad Shojaee, Mohammad Vahidi, Amirali Soheili, Faezeh Ghanimat, Yasaman Keshmiri, Saeed Abdi, et al. COVID-19 and ICU admission associated predictive factors in Iranian patients. Caspian Journal of Internal Medicine, 11(Suppl 1):512, 2020.

[124] Kianoush B Kashani. Hypoxia in COVID-19: sign of severity or cause for poor outcomes. Mayo Clinic Proceedings, 95(6):1094–1096, 2020.

[125] Pawel Grieb, Maciej Swiatkiewicz, Katarzyna Prus, and Konrad Rejdak. Hypoxia may be a determinative factor in COVID-19 progression. Current Research in Pharmacology and Drug Discovery, 2:100030, 2021.

[126] Qingyang Zhong and Jie Peng. Mean platelet volume/platelet count ratio predicts severe pneumonia of COVID-19. Journal of Clinical Laboratory Analysis, 35(1):e23607, 2021.

[127] Xiaofang Zhao, Kun Wang, Peiyuan Zuo, Yuwei Liu, Meng Zhang, Songpu Xie, Hao Zhang, Xinglin Chen, and Chengyun Liu. Early decrease in blood platelet count is associated with poor prognosis in COVID-19 patients–indications for predictive, preventive, and personalized medical approach. EPMA Journal, 11:139–145, 2020.

[128] Fesih Ok, Omer Erdogan, Emrullah Durmus, Serkan Carkci, and Aggul Canik. Predictive values of blood urea nitrogen/creatinine ratio and other routine blood parameters on disease severity and survival of COVID-19 patients. Journal of Medical Virology, 93(2):786–793, 2021.

[129] Rohollah Valizadeh, Azar Baradaran, Azin Mirzazadeh, and LVKS Bhaskar. Coronavirus-nephropathy; renal involvement in COVID-19. Journal of Renal Injury Prevention, 9(2):e18, 2020.

[130] Wenjing Ye, Guoxi Chen, Xiaopan Li, Xing Lan, Chen Ji, Min Hou, D. Zhang, Guangwang Zeng, Yaling Wang, Cheng Xu, et al. Dynamic changes of D-dimer and neutrophil-lymphocyte count ratio as prognostic biomarkers in COVID-19. Respiratory Research, 21(1):1–7, 2020.

[131] Jason Wagner, Andrew DuPont, Scott Larson, Brooks Cash, and Ahmad Farooq. Absolute lymphocyte count is a prognostic marker in COVID-19: A retrospective cohort review. International Journal of Laboratory Hematology, 42(6):761–765, 2020.

[132] Iman Tavassoly. Dynamics of Cell Fate Decision Mediated by the Interplay of Autophagy and Apoptosis in Cancer Cells: Mathematical Modeling and Experimental Observations. Springer, 2015.

[133] Mohammadreza Dorvash, Mohammad Farahmandnia, and Iman Tavassoly. A systems biology roadmap to decode mTOR control system in cancer. Interdisciplinary Sciences: Computational Life Sciences, 12(1):1–11, 2020.

[134] I Tavassoly, J Parmar, AN Shajahan-Haq, R Clarke, William T Baumann, and John J Tyson. Dynamic modeling of the interaction between autophagy and apoptosis in mammalian cells. CPT: Pharmacometrics & Systems Pharmacology, 4(4):263–272, 2015.

[135] Mohammadreza Dorvash, Mohammad Farahmandnia, Pouria Mosaddeghi, Mitra Farahmandnejad, Hosein Saber, Mohammadhossein Khorraminejad-Shirazi, Amir Azadi, and Iman Tavassoly. Dynamic modeling of signal transduction by mTOR complexes in cancer. Journal of Theoretical Biology, 483:109992, 2019.

[136] Iman Tavassoly, Yuan Hu, Shan Zhao, Chiara Mariottini, Aislyn Boran, Yibang Chen, Lisa Li, Rosa E Tolentino, Gomathi Jayaraman, Joseph Goldfarb, et al. Genomic signatures defining responsiveness to allopurinol and combination therapy for lung cancer identified by systems therapeutics analyses. Molecular Oncology, 13(8):1725–1743, 2019.

