## Supplemental Materials for "Decoding Clinical Biomarker Space of COVID-19: Exploring Matrix Factorization-based Feature Selection Methods"

### S Supplementary Material

#### S.1 Results of Classification Metrics

Table S.1: The results of classification metrics of the Random Forest classifier for different feature selection algorithms and different number of selected subset of features $k = \{2, 4, 6, 8, 10\}$.

| Method | Metric | $k = 2$ | $k = 4$ | $k = 6$ | $k = 8$ | $k = 10$ | Average |
|---|---|---|---|---|---|---|---|
| MMFS | ACC | 68.7 | 72.03 | 81.96 | 86.73 | 90.61 | 80 |
| | TPR | 88.12 | 81.98 | 85.71 | 89.79 | 95.72 | 88.26 |
| | TNR | 46.15 | 60.49 | 77.59 | 83.16 | 84.67 | 70.41 |
| | PPV | 65.79 | 71.2 | 81.97 | 86.24 | 87.93 | 78.62 |
| | NPV | 76.33 | 76.75 | 82.68 | 87.67 | 94.5 | 83.58 |
| MPMR | ACC | 69.18 | 74.18 | 81.16 | 85.94 | 91.52 | 80.39 |
| | TPR | 77.01 | 77.13 | 85.7 | 87.73 | 95.91 | 84.69 |
| | TNR | 60.09 | 70.75 | 75.89 | 83.86 | 86.42 | 75.4 |
| | PPV | 69.89 | 76.13 | 80.73 | 86.6 | 89.22 | 80.51 |
| | NPV | 71.77 | 74.14 | 82.57 | 86.11 | 94.9 | 81.89 |
| SGFS | ACC | 68.03 | 74.33 | 81.97 | 86.35 | 90.94 | 80.32 |
| | TPR | 73.13 | 76.47 | 83.89 | 89.47 | 92.98 | 83.18 |
| | TNR | 61.09 | 70.83 | 78.74 | 81.73 | 87.57 | 75.99 |
| | PPV | 70.11 | 77.2 | 83.33 | 86.47 | 91.12 | 81.64 |
| | NPV | 72.01 | 74.56 | 81.94 | 87.74 | 92.34 | 81.71 |
| RMFFS | ACC | 69.15 | 71.59 | 82.94 | 84.33 | 90.92 | 79.78 |
| | TPR | 82.94 | 76.42 | 89.24 | 88.1 | 95.92 | 86.52 |
| | TNR | 53.14 | 65.97 | 75.62 | 79.96 | 85.11 | 71.96 |
| | PPV | 67.69 | 72.57 | 81.25 | 83.64 | 88.39 | 78.7 |
| | NPV | 74.07 | 73.11 | 85.92 | 85.91 | 94.83 | 82.76 |
| SLSDR | ACC | 70.6 | 73.48 | 86.45 | 90.73 | 93.62 | 82.97 |
| | TPR | 86.79 | 83.27 | 90.49 | 95.13 | 95.68 | 90.27 |
| | TNR | 51.78 | 62.11 | 81.76 | 85.61 | 91.22 | 74.49 |
| | PPV | 68.18 | 73.03 | 85.75 | 89.02 | 93.07 | 81.81 |
| | NPV | 77.16 | 77.46 | 88.03 | 93.79 | 94.52 | 86.19 |

#### S.2 Classification Metrics

The classification metrics used in this research are defined as follows. Before describing these metrics, we need to introduce True Positive (TP), True Negative (TN), False Positive (FP), and False Negative (FN) concepts.

1. **Classification Accuracy (ACC):** The classification ACC is defined as the proportion of all records whose labels are correctly predicted. The classification ACC can be obtained as

$$\mathrm{ACC} = \frac{\text{number of TP} + \text{number of TN}}{\text{Total number of records}}.$$

2. **True Positive Rate (TPR):** TPR, aka. sensitivity in medical tests or recall, is defined as

$$\text{TPR} = \frac{\text{number of TP}}{\text{number of TP+number of FN}}.$$

A high TPR implies that the number of FN from a machine learning classifier or a medical diagnostic test is relatively low. Thus, a medical test or a classifier with high TPR is effective for ruling out diseases or a specific condition when the test/classification result is negative. However, since TPR does not take into account false positives, a positive result from a test with high TPR does not assure us that the disease or the condition really exists. In our COVID-19 dataset, a negative prediction by a classifier with high TPR value reliably signifies that a patient with COVID-19 condition will most probably survive.

3. **True Negative Rate (TNR):** TNR (aka. specificity) is defined as

$$\text{TNR} = \frac{\text{number of TN}}{\text{number of TN+number of FP}}.$$

TNR is an important characteristic of many medical diagnostics which evaluates the probability of a negative test given that the patient is healthy. Since TNR has the number of FP in its denominator, a positive test result from a high TNR test implies that the probability of the presence of the disease or condition in the patient can be reliably considered. Since TNR does not include false negatives, a negative result of a high TNR diagnostic test does not necessarily mean the disease or condition does not exist in the patient. In the case of COVID-19 dataset, the positive prediction of a classifier with high TNR can be reliably used to predict that the COVID-19 patient will not survive.

4. **Positive Predictive Value (PPV):** PPV (aka. precision) can be calculated as

$$\text{PPV} = \frac{\text{number of TP}}{\text{number of TP+number of FP}} = \frac{\text{number of TP}}{\text{number of predicted positives}}.$$

PPV evaluates the proportion of positive results in medical tests or machine learning predictions that are actually positives.

5. **Negative Predictive Value (NPV):** NPV can be obtained as

$$\text{NPV} = \frac{\text{number of TN}}{\text{number of TN+number of FN}} = \frac{\text{number of TN}}{\text{number of predicted negatives)}}.$$

NPV calculates what proportion of the negative results in a medical test or the classification by a machine learning model are actually negatives.

The confusion matrix in Figure S.1 demonstrates the relationship between the metrics defined above on the one hand and the ground-truth and predicted class labels (conditions) on the other hand. TPR and TNR basically answer a different type of question than PPV and NPV. While the first two metrics condition on the real outcome (ground-truth labels), the other two metrics condition on the models predictions. In other words, what matters for TPR and TNR is that given the real outcome, what is the probability that the machine learning model had the right classification? However, PPV and NPV are concerned with new observations (or patients in medical cases) where the real outcome is not known until after the event.

In other words, they try to answer the question, given the machine learning model predicted a positive (or negative) condition, what is the probability that the sample or the patient has the positive (or negative) condition?

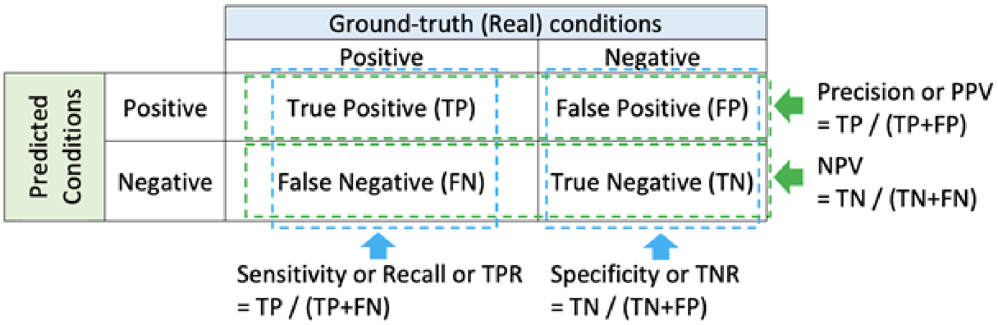


Figure S.1: Confusion matrix to show the relationship between classification metrics on the one hand and the ground-truth and predicted class labels (conditions) on the other hand in a binary classification problem.
